# Household clustering and seasonal genetic variation of *Plasmodium falciparum* at the community-level in The Gambia

**DOI:** 10.1101/2024.08.05.24311344

**Authors:** Marc-Antoine Guery, Sukai Ceesay, Sainabou Drammeh, Fatou K Jaiteh, Umberto d’Alessandro, Teun Bousema, David J Conway, Antoine Claessens

## Abstract

Understanding the genetic diversity and transmission dynamics of *Plasmodium falciparum*, the causative agent of malaria, is crucial for effective control and elimination efforts. In some endemic regions, malaria is highly seasonal with no or little transmission during up to 8 months, yet little is known about how seasonality affects the parasite population genetics. Here we conducted a longitudinal study over 2.5 years on 1516 participants in the Upper River Region of The Gambia. With 425 *P. falciparum* genetic barcodes genotyped from asymptomatic infections, we developed an identity by descent (IBD) based pipeline and validated its accuracy against 199 parasite genomes sequenced from the same isolates. Genetic relatedness between isolates revealed a very low inbreeding level, suggesting continuous recombination among parasites rather than the dominance of specific strains. However, isolates from the same household were six-fold more likely to be genetically related compared to those from other villages, suggesting close transmission links within households. Seasonal variation also influenced parasite genetics, with most differentiation occurring during the transition from the low transmission season to the subsequent high transmission season. Yet chronic infections presented exceptions, including one individual who had a continuous infection by the same parasite genotype for at least 18 months. Our findings highlight the burden of asymptomatic chronic malaria carriers and the importance of characterising the parasite genetic population at the community-level. Most importantly, ‘reactive’ approaches for malaria elimination should not be limited to acute malaria cases but be broadened to households of asymptomatic carriers.

**Author Summary:** Malaria, caused by the parasite *Plasmodium falciparum*, remains a significant health challenge, particularly in regions like The Gambia where transmission is seasonal. Our study explored how seasonal changes impact the genetic diversity and transmission patterns of *P. falciparum* within communities. By tracking 1516 participants over 2.5 years and analyzing the genetic material of the parasites they carried, we discovered that malaria parasites in this region exhibit high genetic diversity, with frequent recombination events rather than dominance by specific strains. Interestingly, we found that parasites from individuals within the same household were much more likely to be genetically related, suggesting close transmission links within households. Seasonal shifts also influenced genetic relationships, with parasite populations becoming more diverse during peak transmission periods. Notably, one individual carried the same parasite strain for over a year without showing symptoms, highlighting the hidden reservoir of chronic malaria infections. Our findings suggest that malaria control strategies should extend beyond treating symptomatic cases to include household members of infected individuals, aiming to disrupt the wider transmission network within communities. This approach could be crucial for advancing malaria elimination efforts in regions with similar transmission dynamics.

## INTRODUCTION

Malaria control and elimination efforts require a detailed understanding of *Plasmodium falciparum* population dynamics, particularly in regions with seasonal transmission. In The Gambia, substantial progress has been made in reducing malaria prevalence in the first decade of 2000. However, the decline has fallen short of meeting the 75% reduction target set for 2025, emphasizing the need for more targeted interventions[1]. In regions characterised by seasonal transmission, malaria clinical cases peak towards the end of the wet season, a pattern mirrored in The Gambia’s transmission cycle. Its climate is characterised by a rainy season from late June to September, followed by a ∼8 month-long dry season, typically without rainfall. Malaria cases are almost exclusively reported from September to December (the high transmission season), while malaria is virtually absent for the remaining 8 months (the low transmission season) [2]. During the high transmission season, malaria disproportionately affects rural, low-income populations in the eastern regions [3–6]. Understanding how the *Plasmodium falciparum* parasite population adapts to these seasonal variations is critical for informing targeted malaria elimination strategies.

Evidence suggests that imported cases play a minor role in maintaining malaria transmission, with the resurgence of malaria attributed to a persistent reservoir of asymptomatic chronic carriers that bridge two transmission seasons separated by several months of low to no transmission [7–10]. Previous studies in The Gambia and neighbouring Senegal demonstrated that individuals can harbor persistent infections across transmission seasons [6,11], with some infections maintaining transmissible gametocytes [12,13]. This reservoir of asymptomatic infections represents a major challenge to malaria control programs Various molecular methods and metrics are now available to characterise the parasite genetic diversity. At the population level, the average COI—the number of distinct parasite genotypes within a host—is often used to infer transmission intensity [14–18]. Genetic relatedness is assessed through Whole Genome Sequencing (WGS) or the cost-effective molecular barcode genotyping, the latter able to identify a unique parasite strain with as few as 24 loci [19]. From pairwise distances between such barcodes, it has been shown that relatedness tends to decrease with time and distance of the sampled infections both at the national level in The Gambia and at the local level in neighbouring Senegal [18,20]. Population-level genetic analyses, particularly using

Identity-by-Descent (IBD), provide a powerful tool for exploring parasite relatedness and recent recombination events. While Fst-based studies are valuable for long-term and large-scale comparisons, IBD allows for the resolution of recent recombination events, shedding light on fine-scale spatio-temporal transmission dynamics [21–23]. Extensive genomic epidemiology studies to date focused on clinical cases, yet such symptomatic cases only represent a minority of all *P. falciparum* infections [24,25]. Thus, relying exclusively on clinical cases may lead to miss a substantial fraction of the actual parasite population diversity.

In this study, we analysed *P. falciparum* genetic diversity in four nearby villages of The Gambia’s Upper River Region through a longitudinal study spanning 2.5 years [11,26]. Using a combination of molecular genotyping and whole genome sequencing, we constructed a high-quality pipeline to assess COI and genetic relatedness at the community level. We aimed to elucidate how seasonal transmission cycles shape parasite population structure, including the stability of COI, the spatio-temporal patterns of IBD, the persistence of drug resistance alleles, and the contribution of asymptomatic chronic carriers to sustained transmission. These findings provide insights into the dynamics of parasite recombination and the genetic adaptations required for persistence in seasonal transmission settings.

## MATERIALS AND METHODS

### Study design and participants

Starting in December 2014, we recruited all residents from two villages (Madina Samako and Njayel, identified respectively with the letters ‘K’ and ‘J’), with two additional villages (Sendebu and Karandaba, identified respectively with the letters ‘P’ and ‘N’) recruited from July 2016, all four villages being in the Upper River Region in The Gambia within 5 km of each other. Active case detection was conducted a total of 11 times over the 2-year period, with each sampling session occurring within a ten-day window. Symptomatic cases that occurred between September and December 2016 were also sampled. More information about the recruited participants can be found in a previous study [26]. In December 2016, 74 asymptomatic *P. falciparum* carriers were sampled monthly for up to 6 months until May 2017, with 42 of them being part of a cohort previously reported [11]. The study protocol was reviewed and approved by the Gambia Government/MRC Joint Ethics Committee (SCC 1476, SCC 1318, L2015.50) and by the London School of Hygiene & Tropical Medicine Ethics Committee (Ref. 10982).

### Sampling and molecular detection of parasites

Details on the *P. falciparum* detection is provided in previous works [11,26]. Briefly, fingerprick blood samples were tested for *P. falciparum* by varATS qPCR. From July 2016 onwards, individuals testing positive for *P. falciparum* were invited to provide an additional 5 to 8 mL venous blood sample that was leucodepleted with cellulose-based columns (MN2100ff) and frozen immediately, as described by MalariaGEN (https://www.malariagen.net/article/online-protocol-leucocyte-depletion-using-mn2100ff-cellulose-columns/). DNA was extracted with QIAgen Miniprep kit following manufacturer procedure.

### Genotyping and genome sequencing

A total of 522 *P*. *falciparum* DNA positive samples, 307 from fingerprick and 215 from venous blood, were processed for genotyping (442 samples) and whole genome sequencing when sufficient parasite DNA was available (331 samples of which 251 were genotyped) as part of the SpotMalaria consortium (Figure 1B). Genotyping was performed by mass-spectrometry based platform from Agena MassArray system. The output consisted of 101 bi-allelic SNPs located on the 14 chromosomes and concatenated into a ‘molecular barcode’ [27], plus six markers of resistance to antimalarials: AAT1 S528L (associated with chloroquine resistance), CRT K76T (associated with chloroquine resistance), DHFR S108N (associated with pyrimethamine resistance), DHPS A437G (associated with sulfadoxine resistance), Kelch13 C580Y (associated with artemisinin resistance) and MDR1 N86Y (associated with chloroquine, amodiaquine, lumefantrine and mefloquine resistance). The 101 SNPs had been picked for their variable allele frequencies within the *P*. *falciparum* population [27]. Whole genome sequencing (Illumina) was performed after a Selective Whole Genome Amplification step [28]. Paired end DNA sequence-reads (150 bp) were aligned to 3D7 reference genome version 3. Variants were called by a script from the MalariaGen consortium using GATK HaplotypeCaller [29,30]. Two drug resistance markers, AAT1 S528L and Kelch13 C580Y were only available in sequencing data and absent from genotyping data. If distinct calls of drug resistance markers were obtained between genotyping and whole genome sequencing data, only the call of whole genome sequencing was considered for the rest of the analysis.

**Figure 1:**
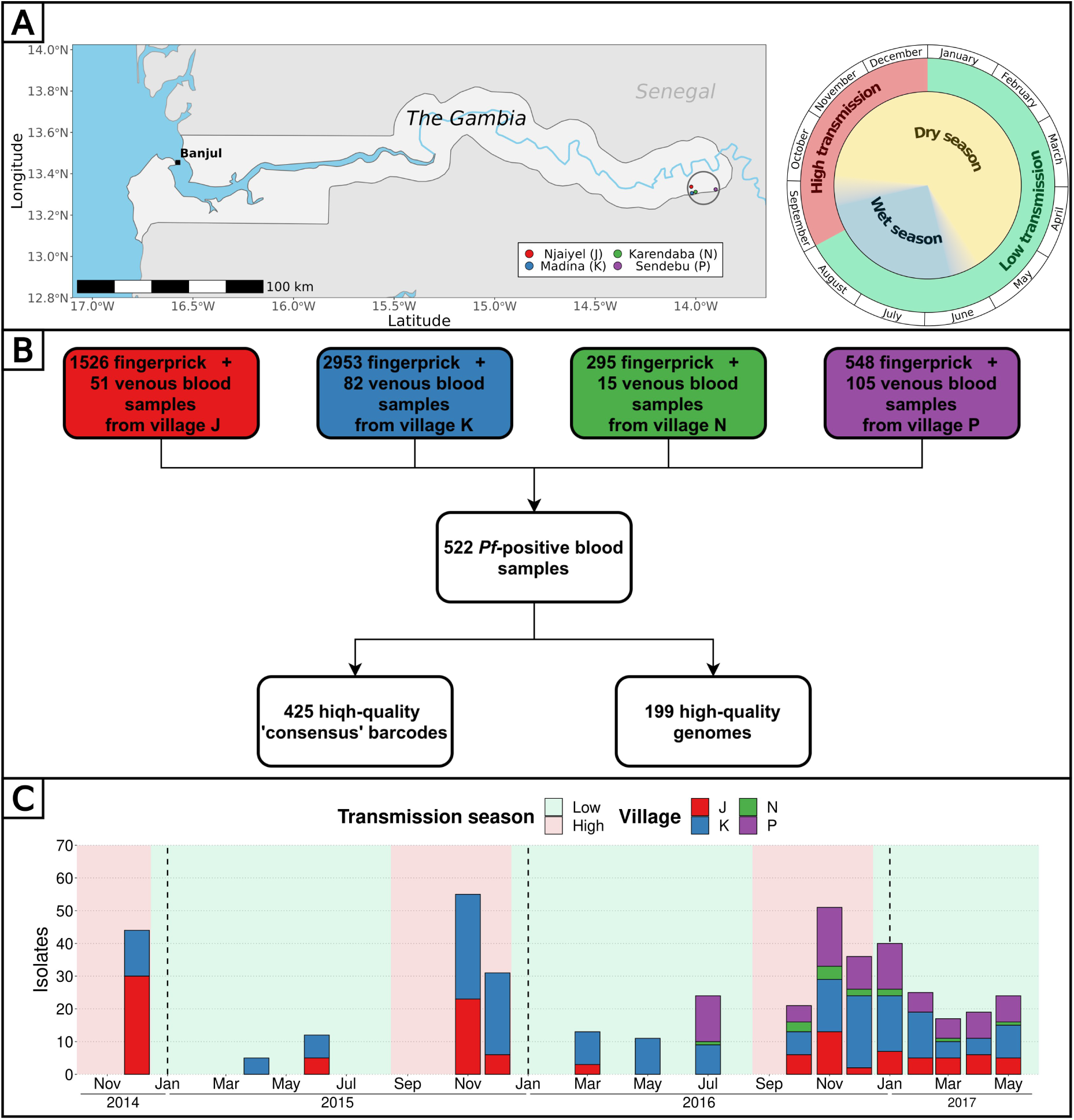
Study design and analysis pipeline. (A) Blood samples from all participants from 4 villages of the Upper River Region of The Gambia were collected up to 16 times over 2.5 years. The peak of clinical malaria cases occurs at the end and right after the rainy season. Made with Natural Earth. (B) Overall, 522 blood samples (307 fingerprick and 215 venous blood) were genotyped and/or whole genome sequenced, resulting in 425 high-quality barcodes and 199 high-quality genomes. Additionally, 6 drug resistance markers were successfully genotyped and/or called from whole genomes, in a total of 438 isolates. (C) High-quality barcodes and genomes were sampled in 4 villages over 16 time points between December 2014 and May 2017.

### Parasite relatedness

To accurately assess the parasite genetic similarity between different sampled infections, we estimated pairwise mean posterior probabilities of Identity-By-Descent (IBD) between genomes or barcodes using hmmIBD, a hidden Markov model-based software relying on meiotic recombination events given a recombination rate of *Plasmodium falciparum* of 13.5 kb/cM [31,32]. To estimate IBD, the model requires pairs of homozygous sites between two samples that are referred to as ‘comparable sites’ in this manuscript. These comparable sites are used by hmmIBD to yield the probability of two samples to be in IBD, which is equivalent to the expected shared fraction of their genomes. With an IBD of more than 0.9, two samples are considered identical, hence describing the same parasite genotype.

When samples are highly polyclonal, their set of homozygous sites tend to be made out of mostly alleles nearly fixed in the population, with a resulting IBD artificially high. Setting a minimal number of comparable sites for which at least one sample possess the minor allele will reduce this bias; these sites will be referred to as ‘informative sites’ in this manuscript.

### Multi-locus genotype barcode data analysis pipeline

We developed a pipeline in R (version 4.3.3) combining Whole Genome Sequencing (WGS) and molecular genotyping data to produce a genetic relatedness network of parasites.

To analyse WGS data formatted in a VCF format, we developed the following pipeline to (Figure S1):

1. Filter out genomic loci for which the population minor allele frequency is inferior to 0.01, with more than 2 alleles identified in the population or that are located outside of the core genome [32]. Out of 1,042,186 initial SNPs, 27,577 (minimal QUAL of 132) were retained.
2. Remove genomes comprising less than 3000 SNPs covered by at least 5 reads.
3. Estimate the proportion of polyclonal samples using the *F*_ws_ metric and heterozygous loci.
4. Format SNPs into a matrix as required by hmmIBD. The matrix was formatted as follow: 0 and 1 were used for the respective allele of each SNP and -1 for mixed and unknown positions (which were then ignored during the IBD estimation). SNP calls were considered mixed if the within-sample Minor Allele Frequency (MAF) was greater than 0.2. The MAF of 0.2 was chosen according to the good agreement between molecular barcodes and genomic barcodes (high number of heterozygous call matches and low number of heterozygous-homozygous call mismatches) (Figure S2).
5. Use the paired IBD values obtained from hmmIBD and build a network of genome relatedness. IBD values were considered unknown between pairs of genomes having less than 100 informative sites (all our pairs had at least 1000 comparable sites).

The second part of the pipeline imputes missing SNPs in the molecular barcode from the WGS data to build a ‘consensus barcode’ (Figure S1):

1. Build a ‘molecular barcode’ out of the initial 101 genotyped SNPs and a ‘genomic barcode’ out of the same 101 SNPs called from high-quality genomes.
2. Remove 12 loci that are absent from all high-quality genomes.
3. Estimate the within-sample Minor Allele Frequency (MAF) to use as a cutoff to consider a locus homozygous when building barcodes out of genomic data. Genomic barcodes are built using different cutoffs of MAF below which a locus is considered homozygous and aligned against molecular barcodes from the same isolates. The cutoff of within-sample MAF of 0.2 showed a high number of heterozygous call matches and low number of heterozygous-homozygous call mismatches, meaning that molecular barcodes and genomic barcodes were in good agreement (Figure S2). This cutoff was retained to build all genomic barcodes. Each pair of molecular and genomic barcodes obtained from the same isolates were aligned. For isolates sampled after May 2016, molecular barcodes are most of the time not matching genomic barcodes for 21 loci (Figure S3). These 21 loci were clustered in one the 4 multiplexes used for molecular genotyping that probably failed to give the proper base calling. As a result, these 21 loci were considered unknown for all the molecular barcodes obtained after May 2016.
4. Using the alignment between molecular and genomic barcodes from the same isolates, replace unknown and mismatched SNP of the molecular barcodes by the SNP of genomic barcodes. These improved molecular barcodes are referred to as ‘consensus barcodes’.
5. Remove consensus barcodes with fewer than 30 SNPs.
6. Estimate the proportion of polyclonal samples using heterozygous loci (described in following section).
7. Format consensus barcodes into a matrix as required by hmmIBD. The matrix was formatted like with genome data.
8. Use the paired IBD values obtained from hmmIBD and build a network of barcode relatedness. IBD values were considered unknown between pairs of barcodes having less than 30 comparable sites and 10 informative sites.

### Complexity of infections

The clonality of each isolate was estimated from whole genome sequenced samples by the *F*_ws_ metric based on allelic frequencies from genomic data [33]. Additionally, the complexity of infections was estimated by the proportion of heterozygous loci (polyclonal if the proportion is above 0.5 % of available sites) in both consensus barcodes and genomes using a within-sample MAF of 0.2.

### Genetic relatedness between groups of multi-locus genotype barcodes

All ‘consensus barcodes’(hereafter referred to as ‘barcodes’) available after December 2016 were excluded from the genetic relatedness analysis as they were obtained exclusively from the 74 individuals selected for their asymptomatic carriage of *P. falciparum* [11]. IBD was used to classify barcode (‘barcode-IBD’) pairs as related (IBD ≥ 0.5) or unrelated (IBD < 0.5). This classification was compared to the one resulting from IBD calculated between pairs of genomes (‘genome-IBD’, considered as gold-standard) using Cohen’s kappa [34]. For pairs of isolates with a different classification between barcode-IBD and genome-IBD, only the genome-IBD classification was retained (57/364 falsely related pairs set as unrelated and 58/18626 falsely unrelated pairs set as related). Additionally, the genome-IBD was used to classify barcodes as related or unrelated when the barcode-IBD was not available (710 pairs).

Barcodes were grouped by their collection date (11 time points from December 2014 to December 2016), sampling location (households or villages) or collection date split by household groups (within the same household, two households of the same village or two households of different villages). Within the same group, the genetic relatedness was estimated by the proportion of related barcodes (IBD ≥ 0.5) over all possible pairs of barcodes. When comparing different groups, the genetic relatedness was estimated by the proportion of related barcodes over all pairs of barcodes of each group, excluding pairs involving barcodes from the same individual. We used a stringent criteria of filtering out pairs of collection dates split by household groups with less than 5 comparisons (25/192 removed pairs). All pairs of villages (10 pairs) and all pairs of dates (66 pairs) contained at least 10 comparisons.

## RESULTS

### Combined barcode and whole genome analysis pipeline

Overall, 5322 fingerprick and 253 venous blood samples were collected from 1516 individuals aged 3 to 85 years in four nearby villages of the Upper River Region of The Gambia between December 2014 and May 2017, as detailed in previous studies (Figure 1A) [11,26]. To characterise the *P. falciparum* population genetic relatedness and identify the impact of antimalarial drugs, we attempted parasite genotyping on 442 isolates (Table S4) and whole genome sequencing on 331 isolates (Table S5) for a combined total of 522 unique isolates over 16 time points (Figure 1B). Whole parasite genomes were successfully sequenced for 199 highly covered (between 3756 and 27516 high quality SNPs) isolates distributed across all 16 time points (Figure S4). Through the concatenation of SNPs from molecular barcode genotyping and whole genome sequencing, we obtained a high-quality ‘consensus’ barcode for 425 isolates comprising on average 65 SNPs (median of 64 SNPs, range 30 to 89 SNPs) (Figure 1C, FigureS5). A detailed description of the pipeline is available (Figure S1).

### Complexity of infection is stable across seasons

The complexity of infection (COI), defined as the number of unique genotypes/genomes within an infected individual, serves as an indicator of parasite strain diversity within the population. We estimated COI using *F*_ws_ values from genomes and the proportion of heterozygous loci from genomes and barcodes. Across these metrics, the proportions of polygenotype isolates were estimated as 40 % (ranging from 22 to 61 % between time points) using *F*_ws_ (80/199), 39 % (range: 22 to 57 %) using heterozygous loci of genomes (78/199) and 34 % (range: 18 to 62 %) using heterozygous loci of barcodes (145/425). As expected, the proportion of heterozygous loci showed a strong negative correlation with *F*_ws_ both for barcodes (R^2^ = 0.83, p-value < 10^-15^) and genomes (R^2^ = 0.92, p-value < 10^-15^), indicating its efficacy as a predictor of COI (Figure S6). Although the proportion of polygenotype isolates fluctuated between time points, no discernible trend was observed between low and high transmission season, suggesting a relatively stable complexity of infections in the population throughout the 2.5-year study duration (Figure S7). At the individual level, we previously showed that the COI was stable in polyclonal infections during the dry season [11].

### Low *P. falciparum* inbreeding at the community-level

To determine the relatedness between isolates and compare malaria parasites from distinct geographical locations and distant times, Identity By Descent (IBD) was calculated pairwise. The reliability of consensus barcodes in identifying genetically related isolates was confirmed through a strong linear correlation between barcode-IBD and genome-IBD, particularly when both were above 0.5 (R^2^ = 0.77, p-value < 10^-15^), with this threshold chosen to distinguish between related (IBD ≥ 0.5) and unrelated (IBD < 0.5) samples for all 425 consensus barcodes (Figure S8). The classifications (related or unrelated) resulting from barcode-IBD values and genome-IBD values (considered as the gold-standard) were in a strong agreement (Cohen’s kappa of 0.839) with high values of specificity (0.997), sensitivity (0.841) and precision (0.843).

The level of inbreeding in the parasite population was characterized from relatedness estimated between barcodes sampled up to December 2016, discarding subsequent samplings in 2017 as they concerned exclusively a cohort of 74 asymptomatic carriers. To avoid sampling bias, only one barcode per continuous infection was retained, resulting in 284 unique barcodes sampled between December 2014 and December 2016. Among 34,326 pairwise comparisons of barcodes, only 435 (1.3 %) demonstrated relatedness with an IBD above 0.5, indicating extensive outcrossing within the parasite population (Figure 2A). Out of the 284 barcodes, 73 (26 %) were identical (IBD ≥ 0.9) to a barcode from another individual, while 140 (49 %) were related (0.5 ≤ IBD < 0.9) to at least one other barcode (Figure 2B). Although clusters of genetically related barcodes may suggest a higher connection within villages (1.6 % of related barcodes) than between them (0.7 % of related barcodes), the average proportion of related isolates was not significantly different (Welch t-test value = 1.42, p-value = 0.24), indicating interconnectedness between villages.

**Figure 2:**
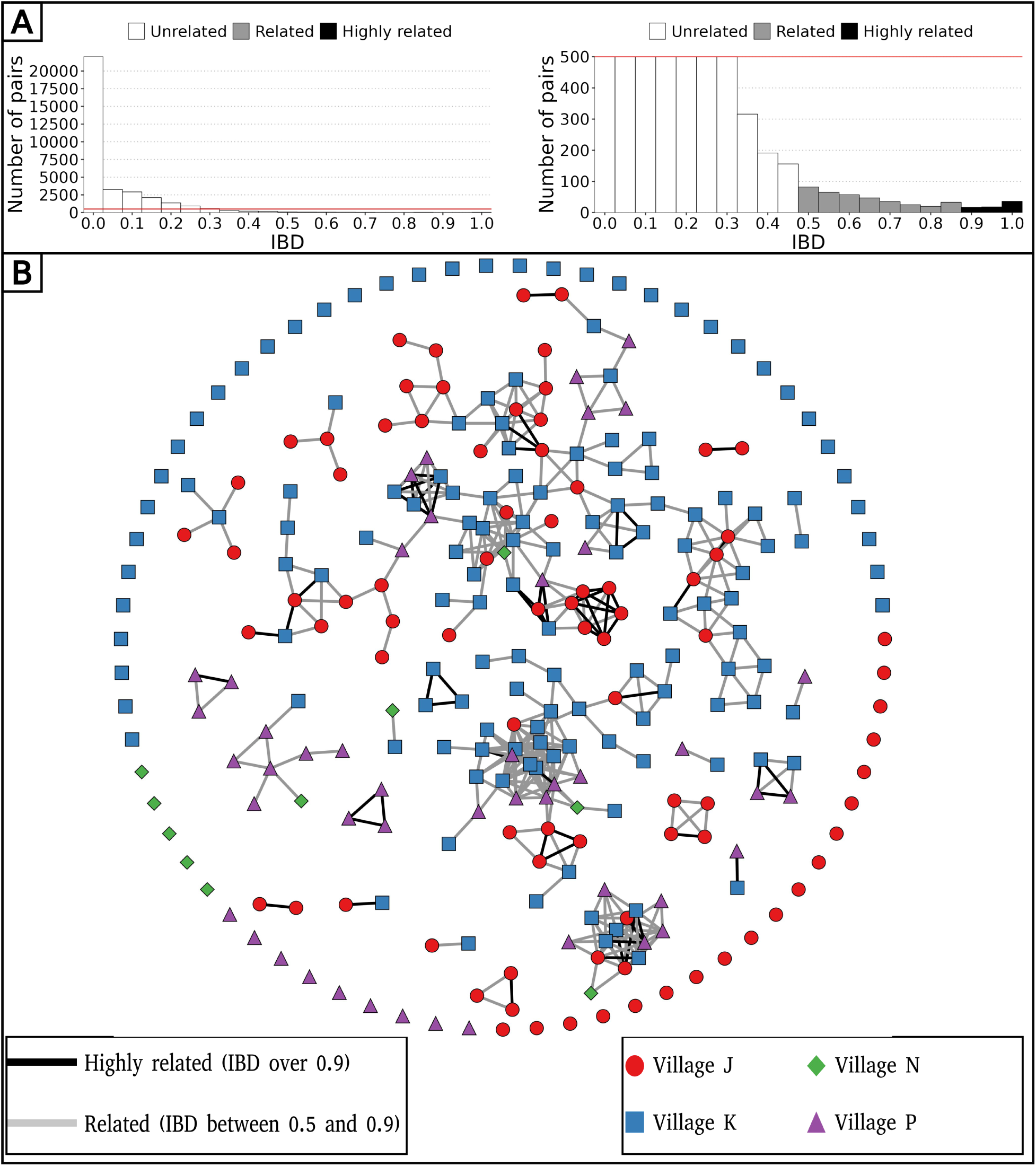
Low parasite inbreeding level in 4 villages in The Gambia inferred from inter-individual genetic relatedness. The genetic relatedness of parasites was assessed from barcodes sampled between December 2014 and December 2016, keeping just one barcode per continuous infection, which resulted in 284 remaining barcodes. (A) Distribution of IBD values between barcodes (left panel), and with a cap at 500 pairs to highlight related barcodes at lower frequency (right panel). (B) Relatedness network of 284 isolates with barcodes represented as nodes and IBD values represented as edges. Barcodes are grouped into clusters using the compound spring embedder layout algorithm from Cytoscape (version 3.10.1).

### Pattern of relatedness between infections is shaped by seasonality

To explore the spatio-temporal relationship between barcodes from distinct individuals, we took advantage of our frequent samplings between December 2014 and December 2016 at the community-level to calculate the percentage of related isolates across six temporal groups, ranging from 0 to 2 months apart to 16 to 24 months apart between sample collections (Figure 3A). Remarkably, at the temporal level, the average proportion of related barcodes within barcodes sampled less than two months apart is 4.7 %. This value is ten-fold higher than the proportion of related barcodes sampled more than 12 months apart equal to 0.30 % (Welch t-test value = 4.72, p-value < 10^-4^). This indicates that recombination between *P. falciparum* isolates breaks down IBD with time.

**Figure 3:**
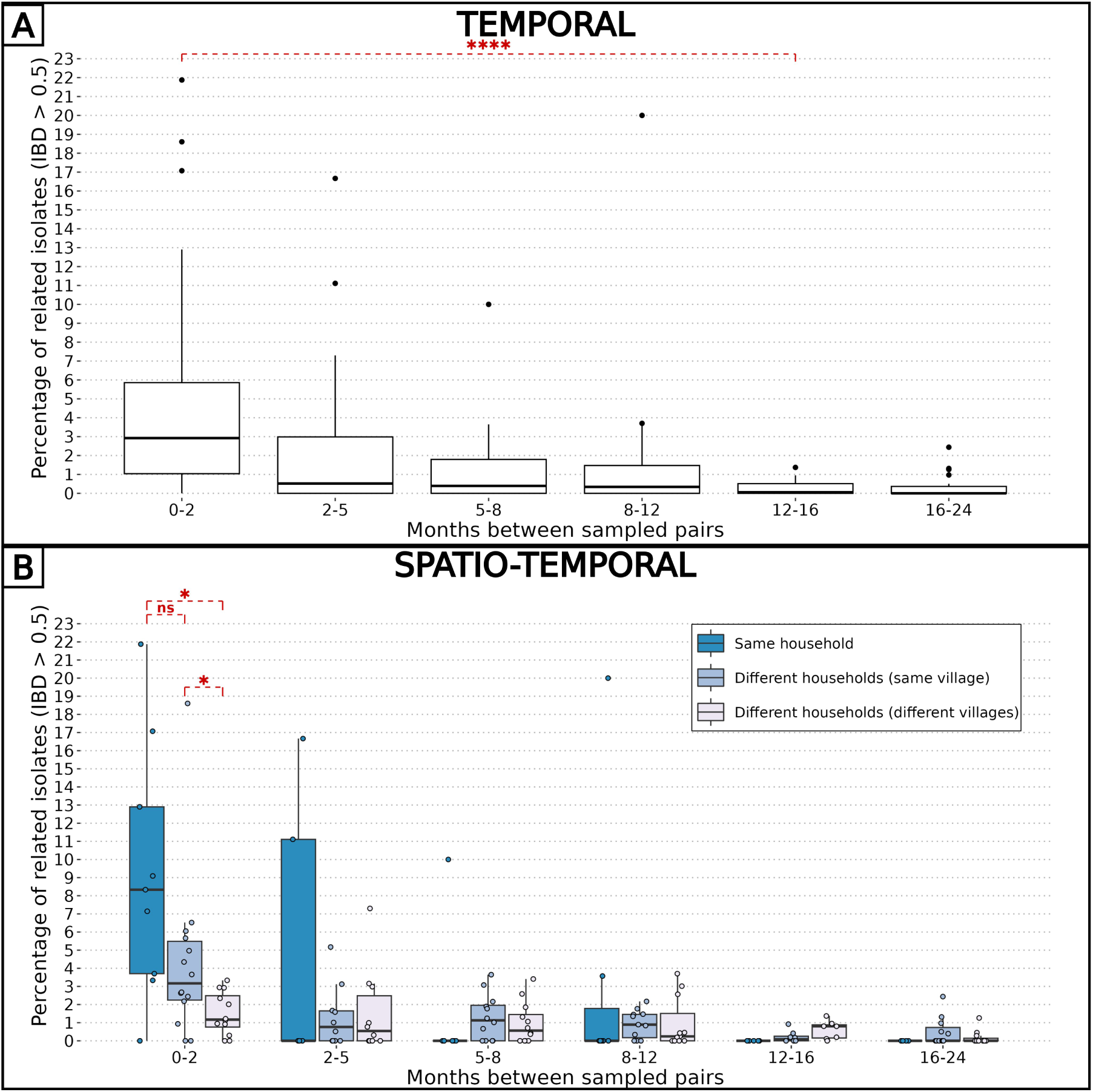
Combined effects of spatial and temporal distances on parasite relatedness. The proportions of related barcodes (IBD ≥ 0.5) between each pair of households are binned into time intervals of various lengths such that the number of observations in each bin is similar. Each box shows the 25-75 % interquartile range of the percentage of related barcodes, with horizontal bar as the median. Groups of related barcodes were compared with Welch t-tests (*: p-value < 0.05, ****: p-value < 0.00005, ns: non-significant). (A) All pairs of isolates were grouped together in the same time interval. (B) Pairs of isolates were grouped by their relative spatial distance.

Similarly, the pairwise IBD was computed for spatial groups: pairs of barcodes from the same household, different households within the same village, and different households across villages (Figure 3B). At the spatial level, barcodes sampled less than 2 months apart and from the same household were six times more related than those sampled from different villages (average proportions of 0.093 and 0.015, Welch t-test value = 3.30, p-value < 0.05). However, when barcodes were sampled more than two months apart, the correlation between genetic relatedness and sampling location disappeared. Altogether, the overall low inbreeding combined with the increased proportion of related isolates within the same household indicate a scenario in which the same infectious mosquito infected two or more individuals living together, or a direct transmission chain between two household members.

The impact of seasonality on the parasite inbreeding level was assessed by comparing the proportion of related barcodes (IBD ≥ 0.5) between groups of collection dates within or between transmission seasons. Barcodes sampled during the high transmission seasons of 2014, 2015 and 2016, as well as the low transmission seasons of 2015 and 2016, were grouped into intraseasonal pairs if they were collected during the same season or interseasonal pairs otherwise. This resulted in 5 groups for intraseasonal pairs and 10 groups for interseasonal pairs, with the most distant groups (high 2014 and high 2016) being 4 seasons apart (Figure 4A). As in Figure 3, pairs of collection dates close in time exhibited greater similarity than more distant collection dates. This suggests a continuous recombination process among all parasites, rather than the transmission of one or more specific strains. To determine the specific time of the year the average genetic relatedness declines, the proportion of related barcodes was compared between pairs of sample collections from the same season or one season apart (Figure 4). Parasites from the ’low to high’ group exhibited the lowest average proportion of related barcodes (average relatedness of 0.006) compared with the ‘within high’ (average relatedness of 0.026, Welch t-test value = 3.14, p-value < 0.01), ‘within low’ (average relatedness of 0.024, Welch t-test value = 2.35, p-value < 0.05) and ‘high to low’ (average relatedness of 0.024, Welch t-test value = 4.87, p-value < 0.001) groups all displaying a 4-fold higher genetic relatedness. Here, most of the parasite differentiation occurs during the transition from the low transmission season to the subsequent high transmission season. This corresponds to the increase in transmission rate at the onset of the high transmission season, with parasite genomes being reshuffled after sexual reproduction in the mosquito. This decrease in relatedness is not observed in the ‘high to low’ group, demonstrating a reduced transmission in the low transmission season.

**Figure 4:**
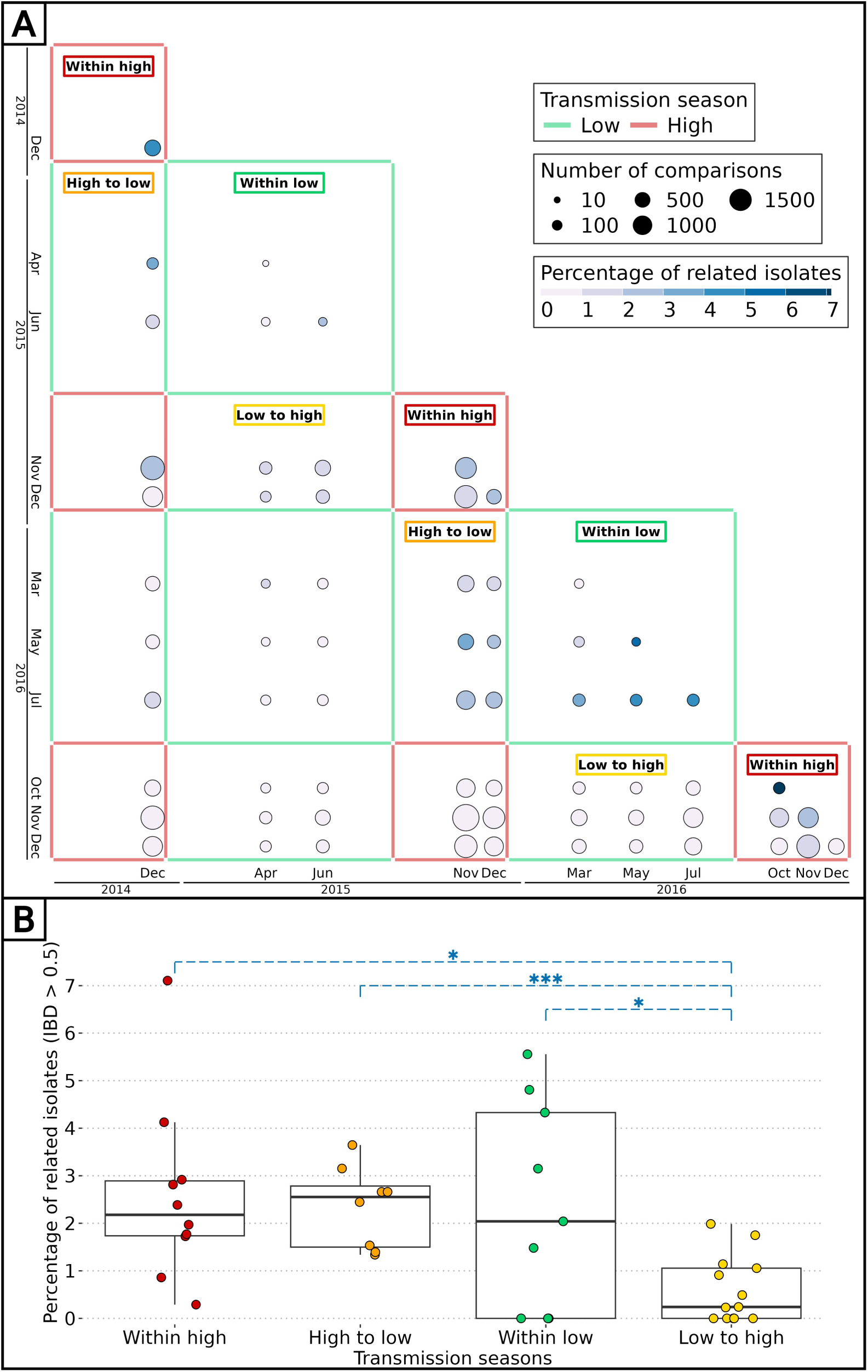
Effect of seasonality on parasite inbreeding level. (A) Proportion of related barcodes (IBD ≥ 0.5) between all sample collections from December 2014 to December 2016. Each square represents a set of pairs of time points within the same seasonal pair. There were 5 successive transmission seasons during the study, hence 15 unique seasonal pairs. Annotations are present for seasonal pairs corresponding to a season compared with itself (‘within high’ and ‘within low’) and to a season compared with the preceding/succeeding one (‘low to high’ and ‘high to low’). (B) Proportion of related barcodes between pairs of sample collections within the same season (‘within high’ and ‘within low’) and one season apart when the high transmission season precedes the low transmission season and conversely (respectively ‘high to low’ and ‘low to high’). Genetic similarities were compared between the ‘low to high’ group and all other groups with Welch t-tests (*: p-value < 0.05, ***: p-value < 0.0005).

### Independence of seasonality and drug resistance markers prevalence

In The Gambia, drug resistant allele frequencies increased dramatically from the late 1980’s until the early 2000’s, then plateaued until 2008 [35]. The rise in clinical malaria cases during the high transmission season implies usage of anti-malaria drugs, particularly among children aged between 0 and 5 years receiving antimalarials (sulfadoxine-pyrimethamine and amodiaquine) monthly during Seasonal Malaria Chemoprevention (SMC), from September to November [36]. The artemisinin-based combination therapy Coartem® (artemether-lumefantrine) is the first-line antimalarial used to treat uncomplicated malaria in The Gambia at the time of the study. We investigated whether the prevalence of drug resistance alleles is influenced by malaria seasonality due to differential selective pressures applied between high (more pressure) and low (less pressure) transmission seasons. To determine the prevalence of resistant haplotypes over time, six drug resistance-related haplotypes were obtained by both molecular genotyping and whole genome sequencing in genes *aat1*, *crt*, *dhfr*, *dhps*, *kelch13* and *mdr1*, leading to a merged total of 438 isolates with drug resistance-related haplotypes. Overall, 89 % of haplotypes called from both molecular genotyping and whole genome sequencing were identical (Figure S9).

As expected, the haplotype Kelch13 C580Y, related to artemisinin resistance, was absent. Overall, the proportion of isolates with resistant alleles was stable over time for AAT1 S258L (0.92, 95 % Wilson’s Confidence Interval: 0.87-0.95), CRT K76T (0.64, 95 % CI: 0.59-0.70), DHFR S108N (0.93, 95 % CI: 0.90-0.95) and MDR1 N86Y (0.12, 95 % CI: 0.09-0.17) (Figure 5). In contrast, the haplotype DHPS A437G appears to decrease in prevalence twice, from the 2015 and 2016 high transmission seasons to the subsequent 2016 and 2017 low transmission seasons. As DHPS A437G haplotype has been associated with resistance to sulfadoxine, its apparent increase in prevalence during high transmission seasons could be resulting from the selective pressure imposed on parasites during SMC. Overall, our data from 2015 to 2017 is very similar to the allele frequency levels from 2008, indicating a potential plateau in the cost-benefit of drug resistance alleles [35].

**Figure 5:**
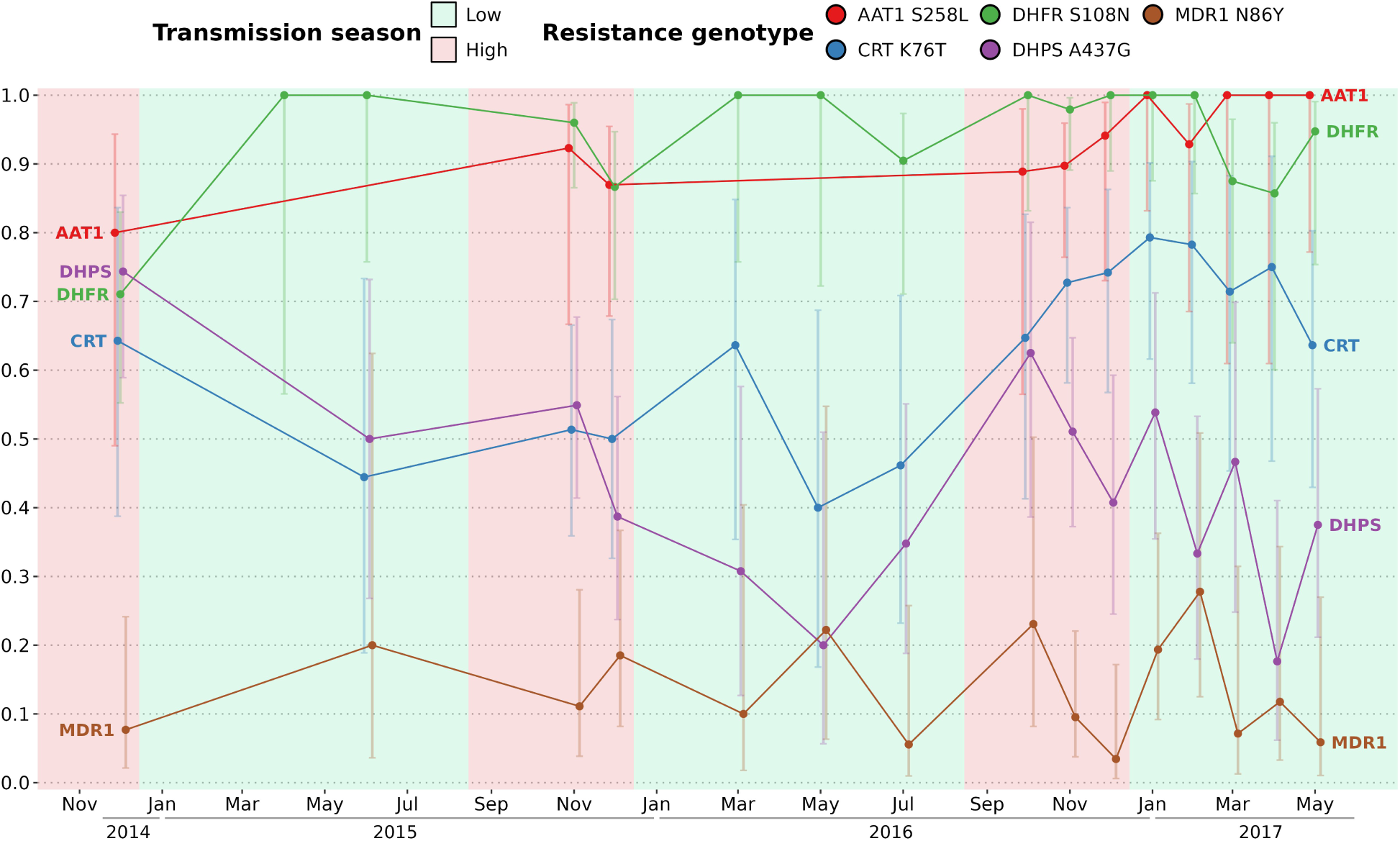
Prevalence of 5 drug resistance-related haplotypes for each time point with at least 10 observations over the study time period. The 5 variants induce non-synonymous changes in AAT1 S528L, CRT K76T, DHFR S108N, DHPS A437G and MDR1 N86Y which are known to reduce the susceptibility to multiple antimalarials. The prevalence of the Kelch13 C580Y mutation was null and not shown here. For all markers except DHPS A437G, the proportions of all variants remained stable between the start and end of the study period. Error bars represent the 95 % Wilson’s confidence intervals.

### *P. falciparum* chronic infections with persisting genotypes

To distinguish reinfections from ‘true’ chronic infections with the same parasite genotype, we measured the minimal duration of infection using IBD values between barcodes obtained from the same individual sampled at different time points (which were not utilized for the spatio-temporal analysis of genetic relatedness). The beginning and end of an infection by the same parasite were attributed to the two most extreme time points separating two identical barcodes (IBD ≥ 0.9). Overall, 32 individuals (20 males and 12 females) were chronically infected with the same dominant *P. falciparum* genotype ranging from one month to one and a half years (Figure 6). Out of the 32 individuals chronically infected, 21 were asymptomatic carriers selected for a monthly sampling between December 2016 and May 2017, rising the chance of observing at least twice the same dominant *P. falciparum* genotype. Gender and age did not significantly influence the duration of infection (Figure S10). Two individuals (24 and 25) were infected in 2017 with two distinct parasite strains, as shown by monthly barcodes alternating between the two strains (for both individuals, barcodes from Feb and Apr, and from Mar and May, are identical). Interestingly, for individual 24, the two strains were genetically related (IBD ≈ 0.5), likely indicating co-infection of sibling strains from the same brood with a fluctuating relative abundance over time. The change in proportion between the two strains over time might be caused by intra-host competition (e.g. one strain, benefiting from immune evasion, is more abundant than the other) or asynchronous developmental stages (one strain is mostly sequestered when the other is mostly circulating and conversely).

**Figure 6:**
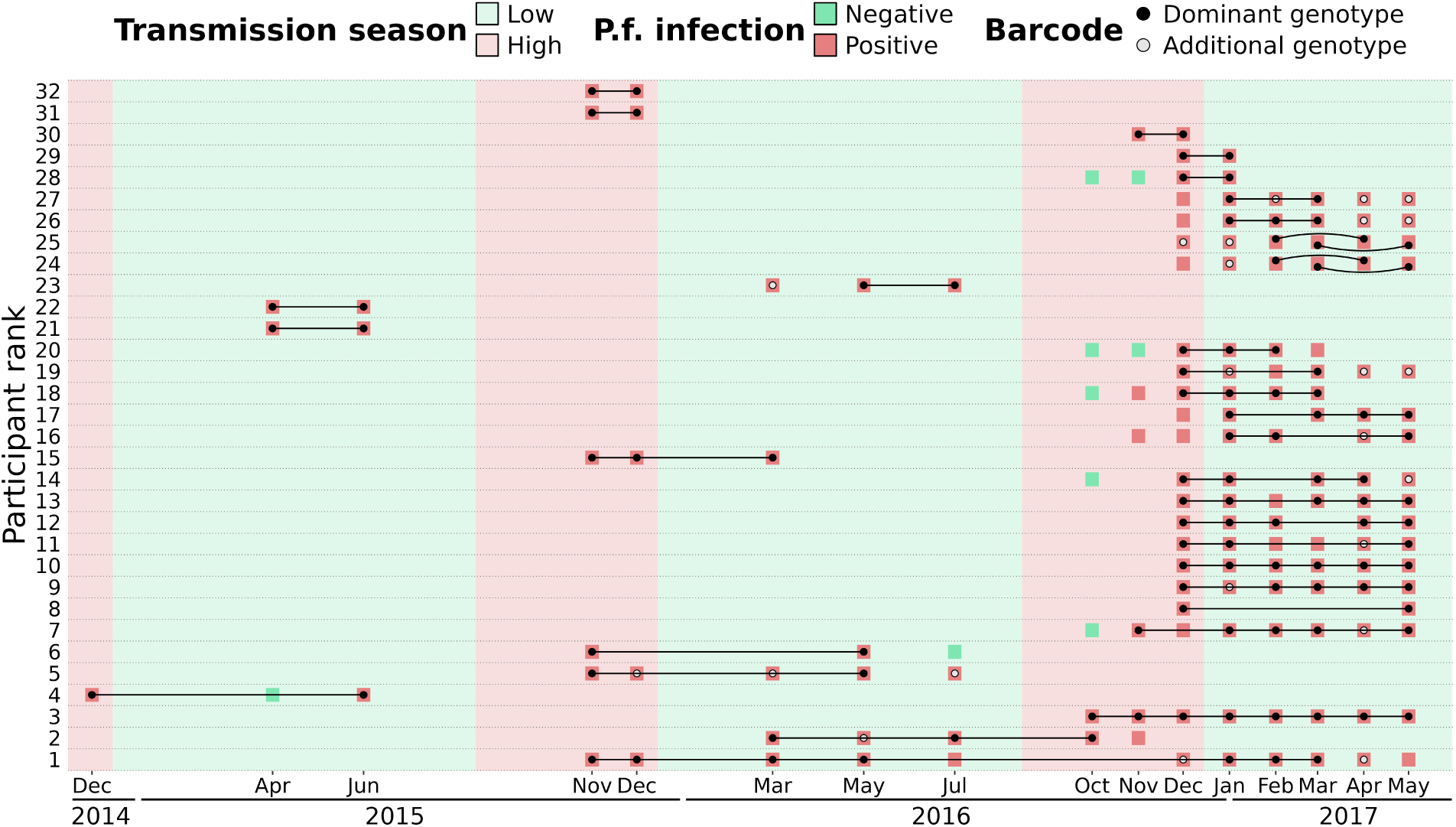
Continuous *P. falciparum* infections with the same dominant genotype. A total of 32 individuals were infected with highly related barcodes (IBD ≥ 0.9) at two or more time points. Individuals are ranked by their duration of continuous infection from the longest to the shortest, with a black line linking identical ‘dominant’ genotypes (black dots) that are the farthest away from each other. Additional genotypes (grey dots) are different (IBD < 0.9) from the dominant genotype but may still be related (IBD ≥ 0.5). For two individuals (ranked 24th and 25th), two different parasite strains were present concurrently, represented by curved lines. Barcodes and *P. falciparum* infection status are shown up to 90 days prior or after the inferred continuous infection.

## DISCUSSION

In anticipation of the pre-elimination phase of malaria, our study aimed to develop robust methods for estimating the complexity of infections (COI) and constructing genetic relatedness networks of *P. falciparum* parasites. By tracking 1,516 participants over 2.5 years, we found that malaria parasites in this region exhibit high genetic diversity, driven by frequent recombination events rather than dominance by specific strains. Notably, parasites from individuals within the same household were significantly more likely to be genetically related, indicating close transmission links within households. Additionally, seasonal changes influenced the genetic relationships among parasites, with populations becoming more diverse during peak transmission periods, reflecting the dynamic nature of malaria transmission in this setting.

Leveraging both barcodes and genomes offers complementary advantages: molecular barcode genotyping provides cost-effective access to genetic information from numerous blood isolates, while whole genomes yields quantitative data on thousands of single nucleotide polymorphisms (SNPs), facilitating comprehensive analyses of parasite diversity, with the added potential to extend analyses to newly discovered haplotypes unidentified at the time of sequencing. Combining both data types validates base calling from molecular genotyping and guides method applicability across the limited positions available in barcodes.

The stable COI over time reflects a constant transmission intensity [16–18,37]. Such stable transmission in this area of The Gambia, reported already elsewhere, contrasts with the overall decrease of malaria transmission across the country, highlighting the heterogeneity malaria within The Gambia [6,26]. Our findings underscore the importance of understanding local malaria dynamics amidst broader regional trends.

With Identity by Descent (IBD), we evaluated parasite relatedness between genomes and between barcodes. Although using more SNPs (e.g. more than 200 SNPs) is ideal to reach an accurate relatedness, *in silico* data suggests that the accuracy is only marginally improved with 96 SNPs or more [38]. Furthermore, we showed that for IBD values above 0.5, the correlation between genome-and barcode-IBD was very strong. Finally, the qualitative aspect of our approach, with isolates either related (IBD ≥ 0.5) or unrelated (IBD < 0.5) enables to compensate for the lower accuracy of barcode-IBD.

Overall, 26 % of unique parasite strains were found in two or more individuals and around 28 % of isolates were polyclonal according to the proportion of heterozygous loci of barcodes. This seemingly contrasts with findings from Thiès, in neighbouring Senegal, where parasite haplotype diversity is low, the proportion of shared genetic types between isolates is 35 %, with less than 10% of polyclonal infections, suggesting self-mating transmission and low outcrossing levels [9,10,39]. However, the key difference of our approach was the active detection of asymptomatic infections, as opposed to the large number of studies sequencing parasites collected from clinical cases. Our results argue for active case detection as a necessary step to comprehensively characterise the parasite genetic diversity.

One limitation of genetic epidemiology studies such as this one, is the necessity to exclude IBD values obtained between isolates with high level of polyclonality and thus not enough informative loci. One important remaining challenge is to incorporate highly polyclonal infections in the network of genetic relatedness of parasites, while the proportion of complex infections tends to increase with malaria transmission intensity [40]. Achieving this goal is difficult as it requires whole genome sequencing followed by a deconvolution tool such as DEploid, with the caveat that the different strains are not too related or too rare [41,42]. A second limitation to our study is that we cannot exclude that imported malaria cases might have an effect on our measured inbreeding levels, enough though previous findings tend to indicate a limited effect of imported cases on maintaining transmission [7–10]. Future studies will require recruiting a larger number of participants with more frequent samplings.

A follow-up study carried out in rural villages of eastern Gambia between 2012 and 2016 reported that roughly half of asymptomatic infected individuals at the end of the wet season were still infected at the end of the dry season [6]. We previously reported a similar rate [11]. We showed that parasites collected during the dry season share significantly more genetic similarity with those from the previous wet season than those from the next wet season. Average relatedness quickly declined with the temporal distance between samplings (10-fold lower after one year) probably due to active genetic recombinations in the population leading to fewer fragments in IBD over time. A study conducted in Colombia (with a 10-fold lower malaria prevalence than in The Gambia) showed that a 10-fold decrease in genetic similarity was reached in 9 years while we observed the same decrease after just one year in our study [37,43].

Most infections were sub-microscopic, which is typically observed when prevalence falls below 20 % [26,44,45]. These low-density infections that may persist for months are typically asymptomatic unless the host immune system is compromised [46]. In this study, the infection with the longest duration was in an asymptomatic individual aged between 5 and 9 years in which it persisted as the same parasite strain for one and a half year. School-age children contribute the most the malaria reservoir through their higher carriage of asymptomatic infections and their ability to effectively infect mosquitoes [6,12,47,48].

In the late 20th century, when the malaria burden in The Gambia was much higher, clinical cases from the same household were more likely to be caused by the same mosquito bite [49]. More recently, Ngwa *et al*. showed that parasite genetic similarity during the 2013 transmission season was inversely correlated with each spatial and temporal distances in The Gambia [20]. We also found that parasites sampled in neighbouring households are more genetically similar but only when sampled less than three months apart. Similarly, two other studies observed parasite strains with varying levels of spatio-temporal propagation in Thiès, Senegal, suggesting that some parasites are more actively transmitted than others [9,18]. These results add evidence that anti-malarial strategies should target all members of a household with an infected individual. Such “reactive” strategies include treatment of a malaria case and all its household members (without testing) or testing all household members and treating if necessary. The impact of reactive strategies on malaria transmission is at best very limited [50–53]. Based on the sheer size of the

*P. falciparum* asymptomatic reservoir, with parasitaemias typically below microscopy or RDT detection level [26], it is not surprising that targeting clinical cases and their immediate families is not sufficient to break the transmission. For a malaria elimination campaign, based on our insights and previous research [54] on asymptomatic infections in The Gambia at the community level, we argue for mass detection with a highly sensitive method such as qPCR, followed by treatment of *P. falciparum* positive cases and all their household members.

## Data availability

The barcode data analysis pipeline can be found at https://github.com/marcguery/malaria-barcodes-genomes-gambia.

Supplementary tables are in ‘Barcode_SuppTables.xlsx’:

- Table S1: Gender, age and households of participants.
- Table S2: Sampling, *falciparum*-malaria test results and treatment.
- Table S3: Locations of genotyped SNPs and their rank in barcodes.
- Table S4: Molecular barcode genotyping raw data and statistics.
- Table S5: Genome sequencing accession identifiers and statistics.
- Table S6: Complexity of infection metrics from barcodes.
- Table S7: Complexity of infection metrics from genomes.
- Table S8: Drug resistance markers genotyped from barcodes.
- Table S9: Drug resistance markers genotyped from genomes.
- Table S10: Duration of continuous infection estimated from barcode relatedness.

This publication uses data from the MalariaGEN SpotMalaria project as described in ’Jacob CG et al.; Genetic surveillance in the Greater Mekong Subregion and South Asia to support malaria control and elimination; eLife 2021;10:e62997 DOI: 10.7554/eLife.62997’ [27]. The project is coordinated by the MalariaGEN Resource Centre with funding from Wellcome (206194, 090770).

## Data Availability

All data produced in the present work are contained in the manuscript and supplementary material.

## Acknowledgements

The authors would like to thank the staff of MalariaGEN, Wellcome Sanger Institute Sample Management, Genotyping, Sequencing and Informatics teams for their contribution. We thank Michael Fontaine, Franck Prugnolle and Virginie Rougeron for insightful comments on data analysis.

## Author contributions

Conceptualization by AC, TB, UDA and DC; field work and sample processing by SC, SD, FJ. Data analysis by MAG and AC. Writing of original draft by MAG and AC. Reviewing and editing of manuscript by all authors. Funding acquisition by AC and TB.

## Supplementary figures

**Figure S1:**
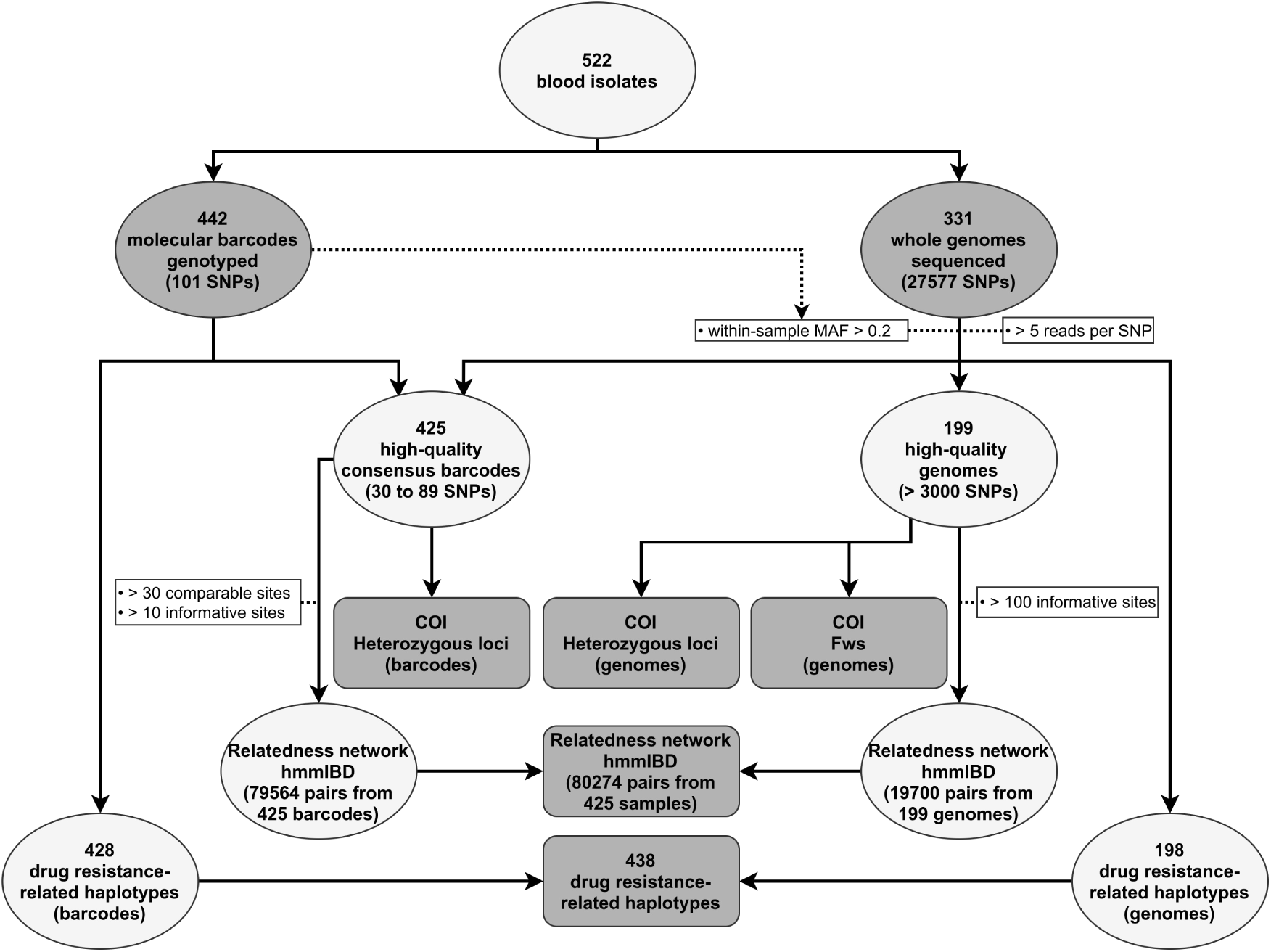
Steps of the combined barcode-and genome-analysis pipeline using 522 *P. falciparum*-positive blood isolates.

**Figure S2:**
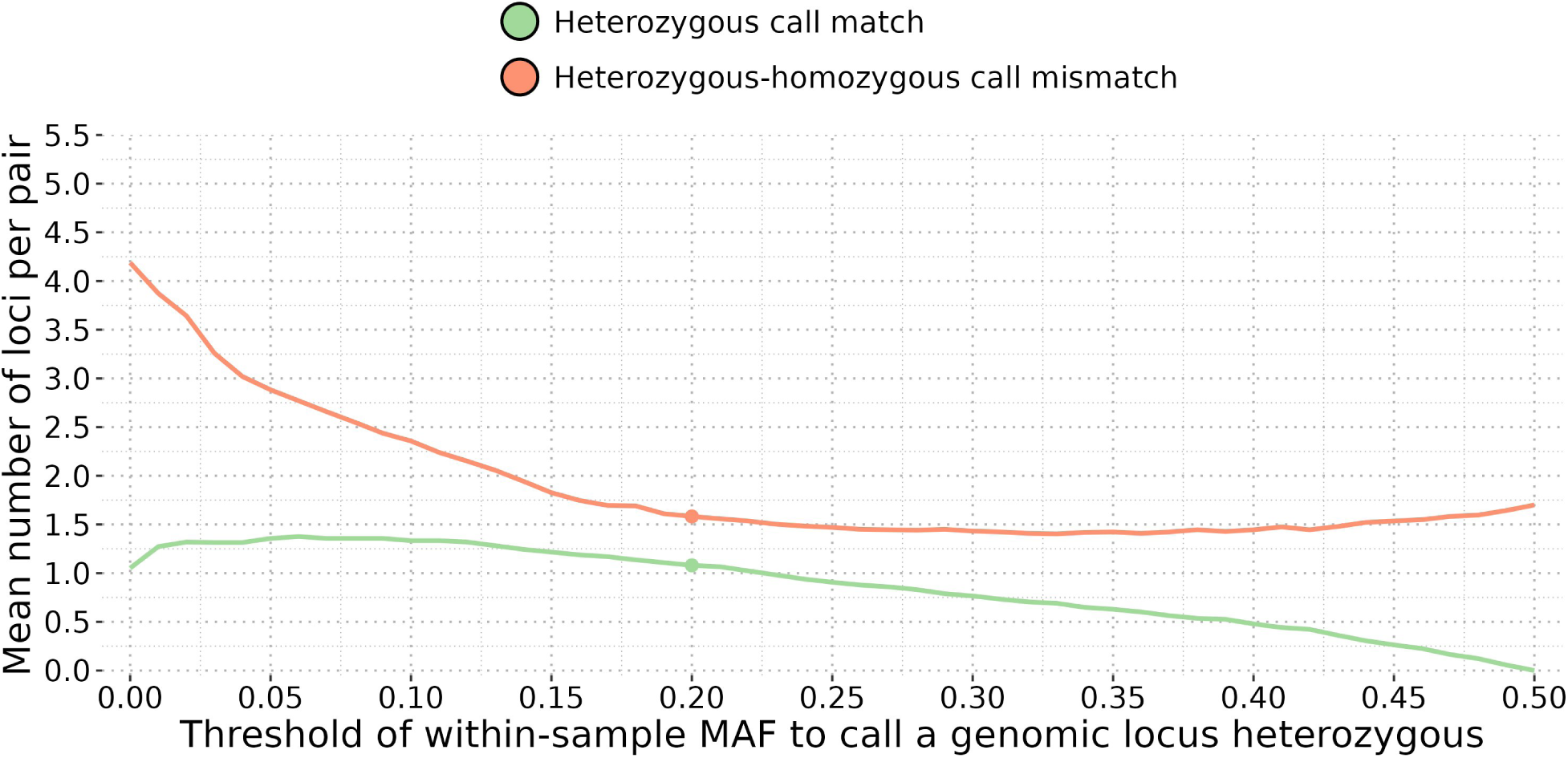
Comparison of heterozygous locus calls between each pair of molecular barcode (obtained by genotyping) and their corresponding genomic barcode (built from allelic frequencies). Genome loci were considered heterozygous if their within-sample Minor Allele Frequencies (MAF) were above a threshold ranging from 0 to 0.5. The threshold of MAF of 0.2 was finally retained because it yields a high number of matches and a low number of mismatches of heterozygous locus call, making the genomic barcodes as close as possible to their molecular counterparts.

**Figure S3:**
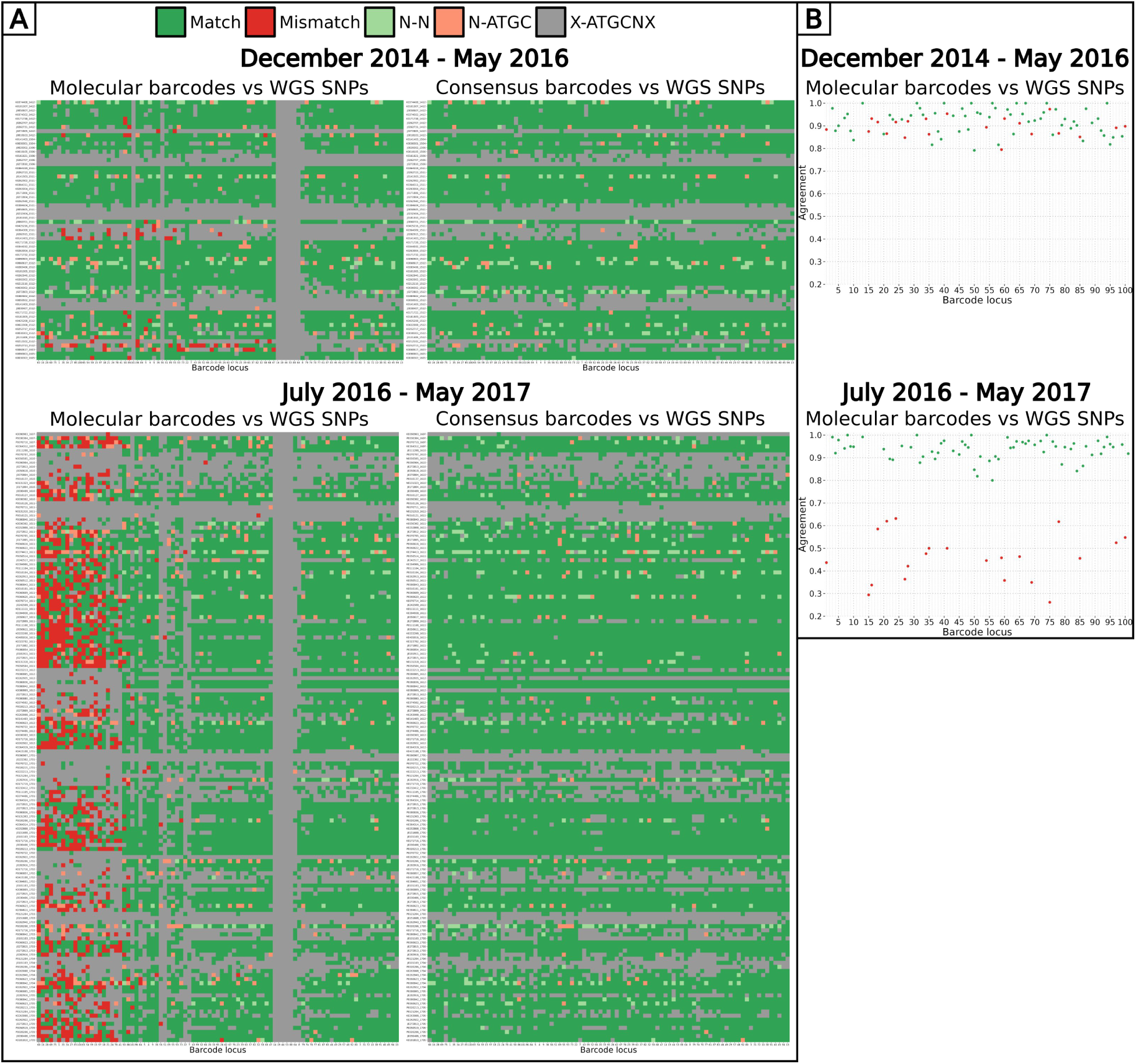
Comparison of molecular barcodes loci (built from genotyped SNPs) and consensus barcodes loci (combining molecular and WGS loci) with WGS loci for two groups of samples clustered by their collection dates. (A) For each sample with both a barcode and a genome, pairs of corresponding loci are colour-coded as ‘matching’ (green), ‘mismatching’ (red), ‘heterozygous call matching’ (light green), ‘heterozygous-homozygous call mismatching’ (light red) or ‘incomparable when at least one call is unknown’ (grey). Barcode loci are sorted based on their agreement with WGS loci (proportion of matching loci over all comparable loci) from low to high.WGS loci were considered heterozygous if the within sample MAF was above 0.2 (Figure S2). (B) Average agreement between molecular barcode loci and WGS loci coloured by their average agreement in samples collected after May 2016 (< 0.8 coloured in red, ≥ 0.8 coloured in green). Barcode loci are sorted by their chromosomal location. Overall, loci match between the two methods for samples collected up to May 2016 (all loci have an agreement above 0.79). However, 21 genotyped loci show a consistent mismatch (all loci have an agreement below 0.7) with WGS loci for all samples collected after May 2016. These 21 genotyped loci were incorrectly genotyped because of a failure of one of the multiplexes during the genotyping procedure. To increase the number of available loci of barcodes, all unknown and incorrectly genotyped loci were replaced by WGS loci, resulting in a ‘consensus barcode’.

**Figure S4:**
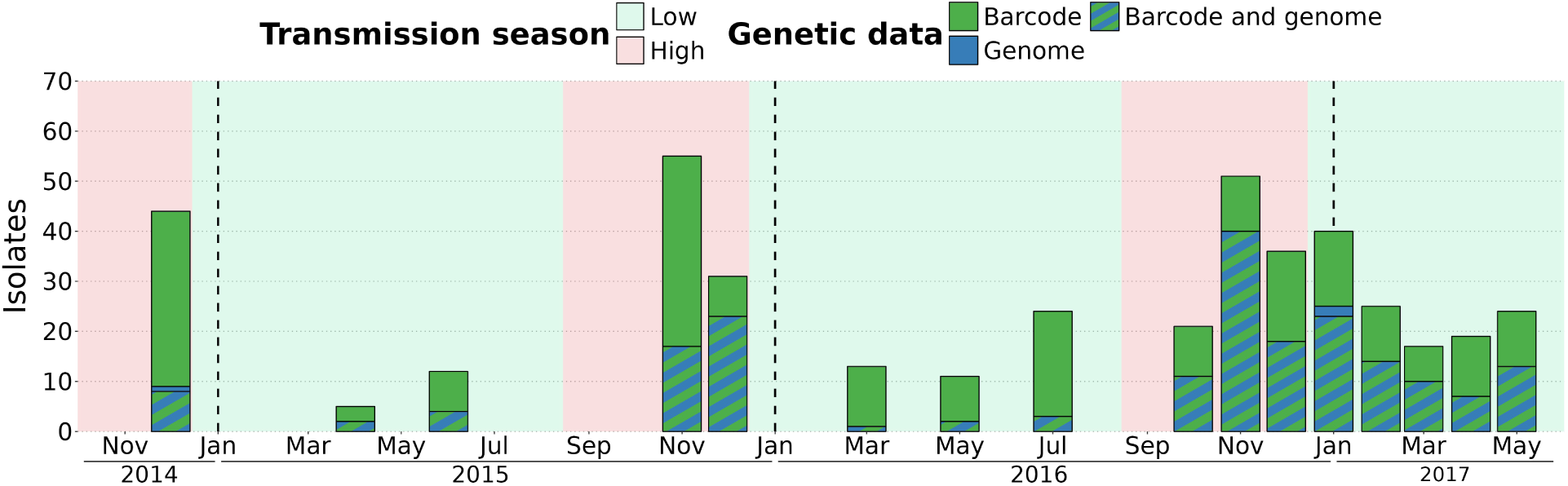
Number of high-quality barcodes (425 isolates) and genomes (199 isolates) successfully sequenced over 16 time points between December 2014 and May 2017.

**Figure S5:**
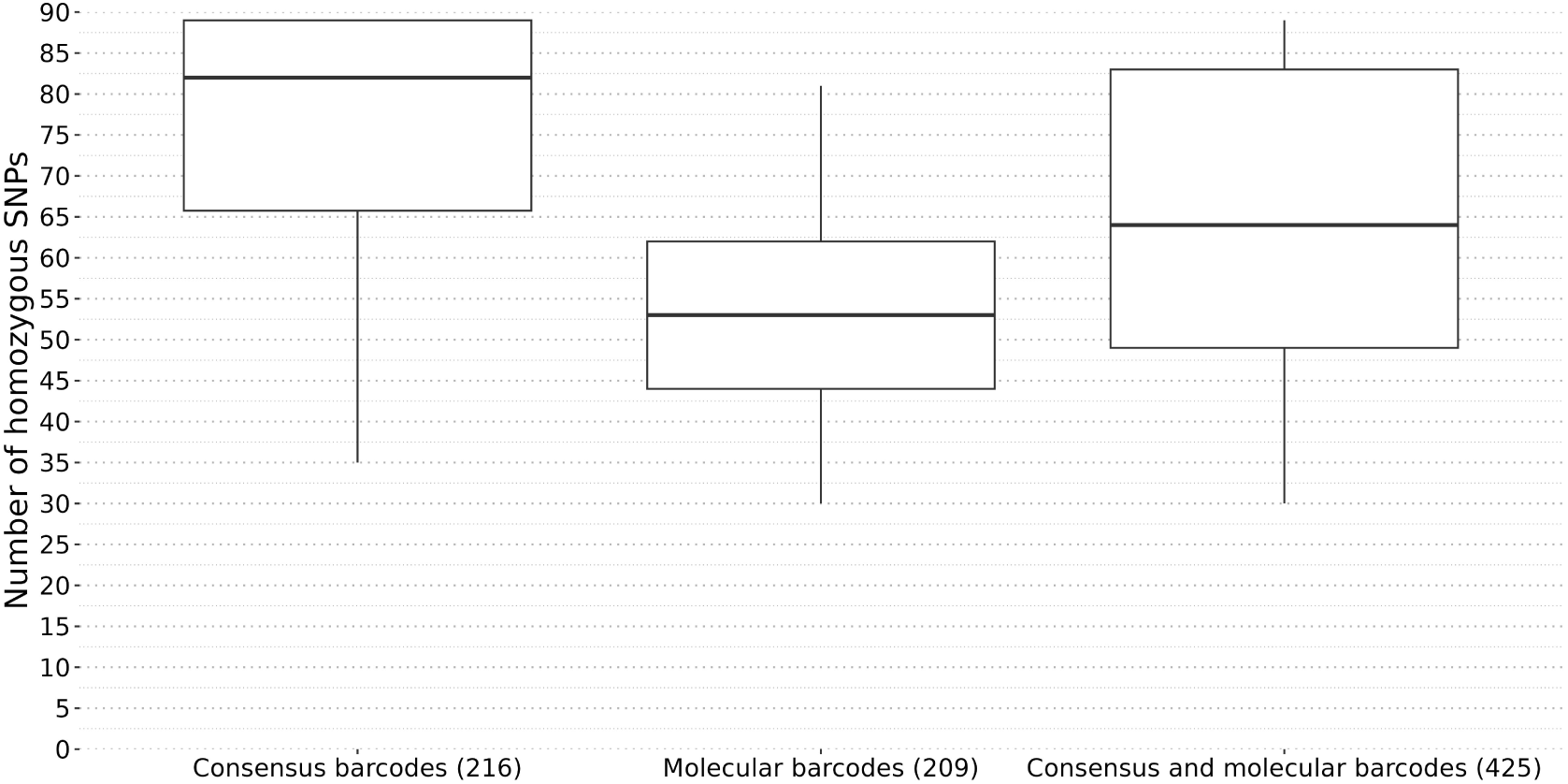
Number of homozygous SNPs per barcode. Among 425 barcodes, 216 were improved using corresponding genomic calls (named ‘consensus barcodes’ on this figure), resulting in an increase in the number of homozygous SNPs (average: 76 SNPs, median: 82 SNPs). The remaining 209 molecular barcodes had a lower number of homozygous SNPs (average: 54 SNPs, median: 53 SNPs). Overall, the 425 consensus and molecular barcodes (referred to as ’consensus barcodes’ in the main text) had an average of 65 SNPs and a median of 64 SNPs.

**Figure S6:**
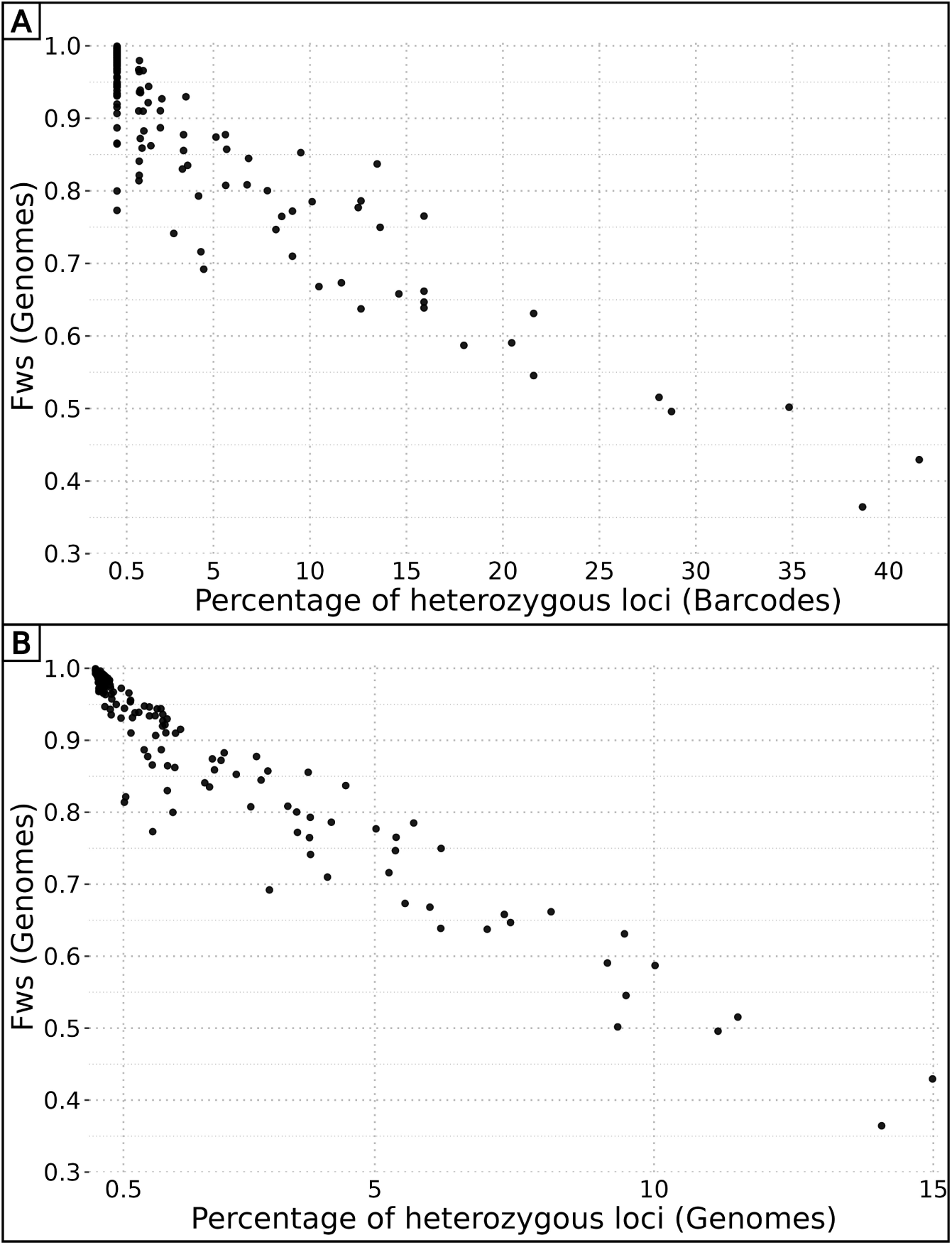
Correlation between *F*_ws_ values and percentage of heterozygous loci. (A) The percentage of heterozygous loci in barcodes ranges from 0 to more than 40 %, highlighting their high variability among *P. falciparum* strains. The percentage of heterozygous loci shows a strong negative correlation with their corresponding *F*_ws_ value (R^2^ = 0.83, p-value < 10^-15^). Considering isolates with *F*_ws_ values available, only 15 % (20/132) of monoclonal isolates (heterozygous site proportion below 0.005) have a *F*_ws_ value below 0.95 and 6 % (4/64) of polyclonal isolates (heterozygous site proportion above 0.005) have a *F*_ws_ value above 0.95. (B) The percentage of heterozygous loci in genomes range from 0 to less than 15 %, which is expected as the 27,577 SNPs had various population-level MAF. The percentage of heterozygous loci shows a strong negative correlation with their corresponding *F*_ws_ value (R^2^ = 0.92, p-value < 10^-15^). Computing the proportion of heterozygous loci in both genome and barcode data (84 SNPs) enables to accurately estimate the level of clonality of an isolate.

**Figure S7:**
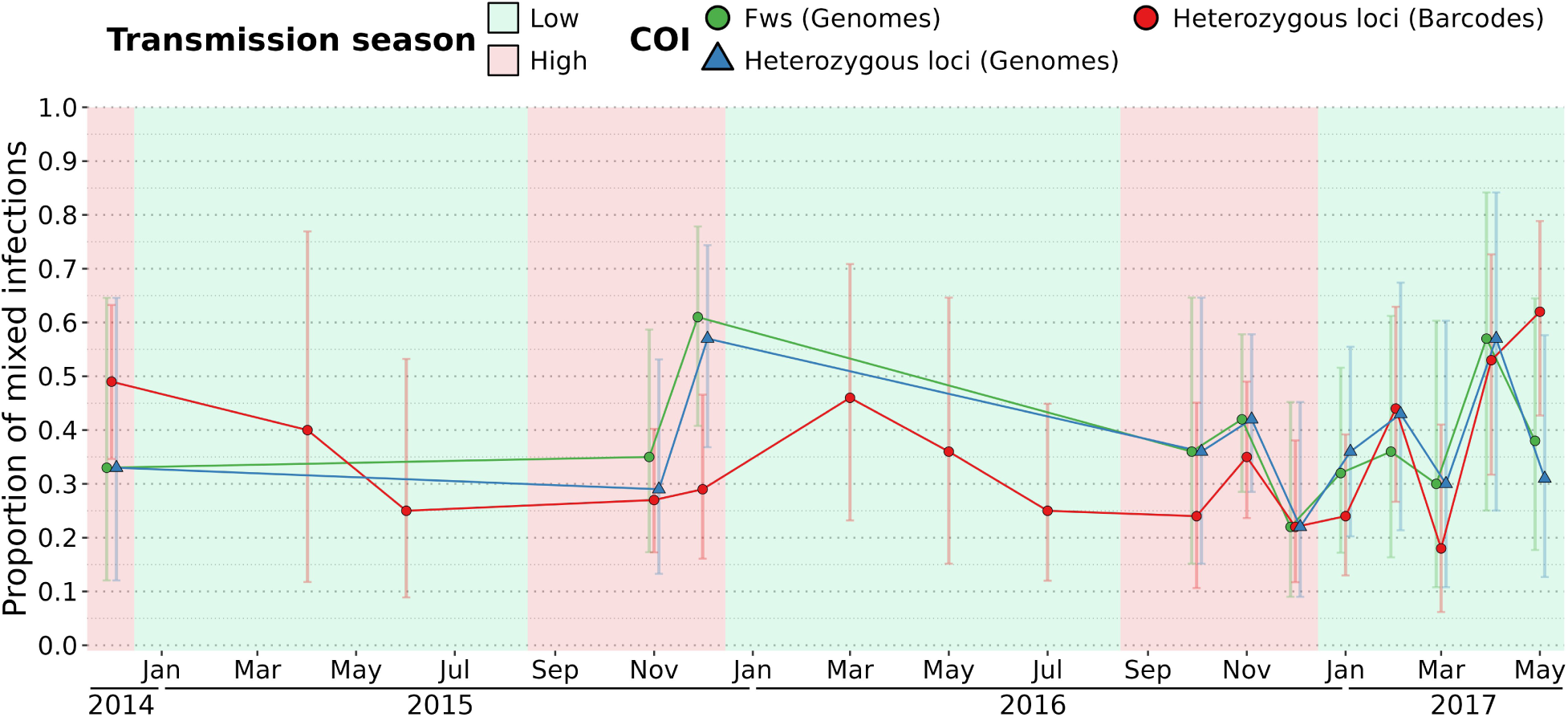
Proportion of polyclonal isolates over time estimated by *F*_ws_ ( *F*_ws_ < 0.95) and the proportion of heterozygous loci (more than 0.5 % of available sites) on barcodes and genomes. The percentage of polyclonal isolates is stable over time for all methods (around 40 % for *F*_ws_, 39 % using heterozygous loci of genomes, 34 % using heterozygous loci of barcodes). Error bars represent the 95 % Wilson’s confidence intervals.

**Figure S8:**
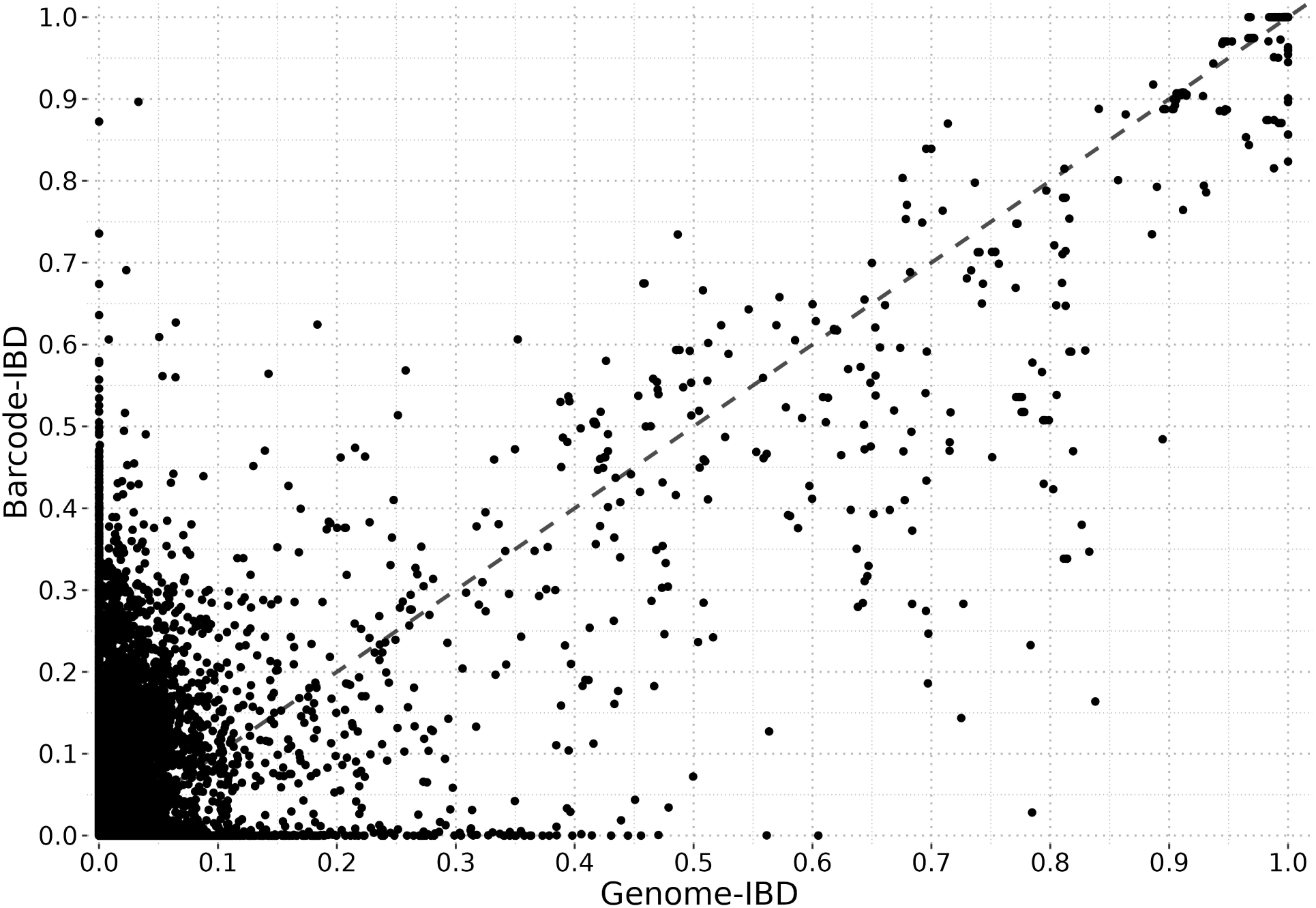
IBD calculated from consensus barcodes highly correlates with IBD from genomes. Identity By Descent was calculated pairwise between the 425 high-quality consensus barcodes (‘barcode-IBD’) with at least 30 comparable sites and 10 informative positions (79564/90100 pairs remaining) and between the 199 high-quality genomes (‘genome-IBD’) with at least 100 informative positions (19700/19701 pairs remaining). The accuracy of the classification of related (IBD ≥ 0.5) and unrelated (IBD < 0.5) isolates from barcode-IBD was assessed using genome-IBD as the gold standard. Overall, the barcode-IBD classification was in a strong agreement with the genome-IBD classification (Cohen’s kappa of 0.839) and had high values of specificity (0.997), sensitivity (0.841) and precision (0.843). Among the 364 pairs of samples with a barcode-IBD above 0.5, 314 were indeed related (genome-IBD above 0.5) while 50 were actually not related (genome-IBD below 0.5), although 24 showed genome-IBD value between 0.4 to 0.5. Regarding the 18626 unrelated pairs according to barcode-IBD classification, only 58 were called as related by the genome-IBD classification.

**Figure S9:**
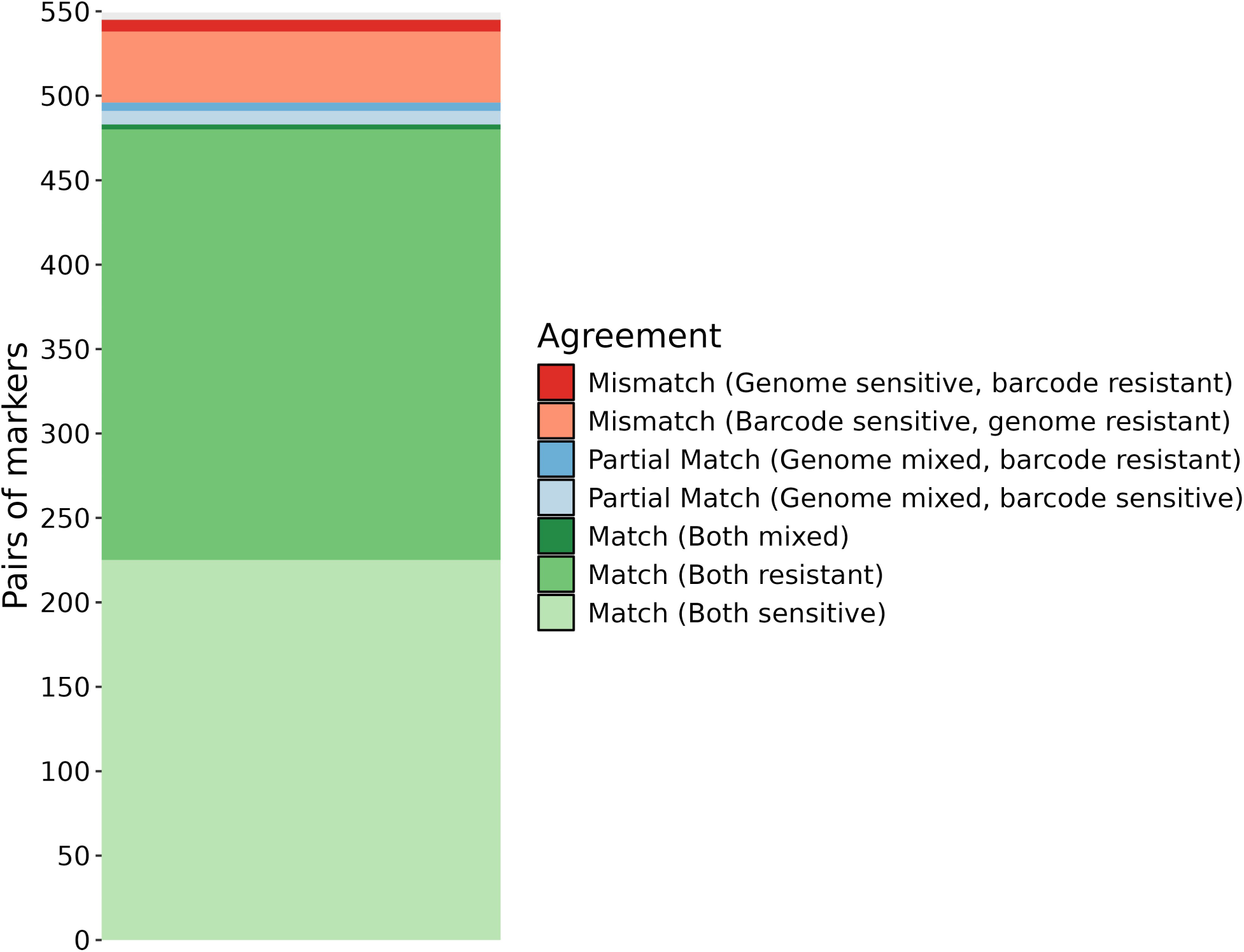
Agreement between pairs of haplotypes obtained both by molecular genotyping (‘barcodes’) and from Whole Genome Sequence calling (‘genomes’). Six drug resistance markers were screened, AAT1 S528L, CRT K76T, DHFR S108N, DHPS A437G, Kelch13 C580Y and MDR1 N86Y. Drug resistance haplotypes were obtained in 428 barcodes (1261 haplotypes) and 198 genomes (1000 haplotypes) for a merged total of 438 isolates (1716 haplotypes). The vast majority (89 %) of the 545 pairs of haplotypes obtained with the two methods are identical, which indicates that both methods are accurate. While there are multiple partial matches (one haplotype is mixed according to a method and either sensitive or resistant by the other) with mixed haplotypes called from genomes, no partial match with mixed haplotypes called from barcodes were found, showing that whole genome sequencing is more sensitive than molecular genotyping.

**Figure S10:**
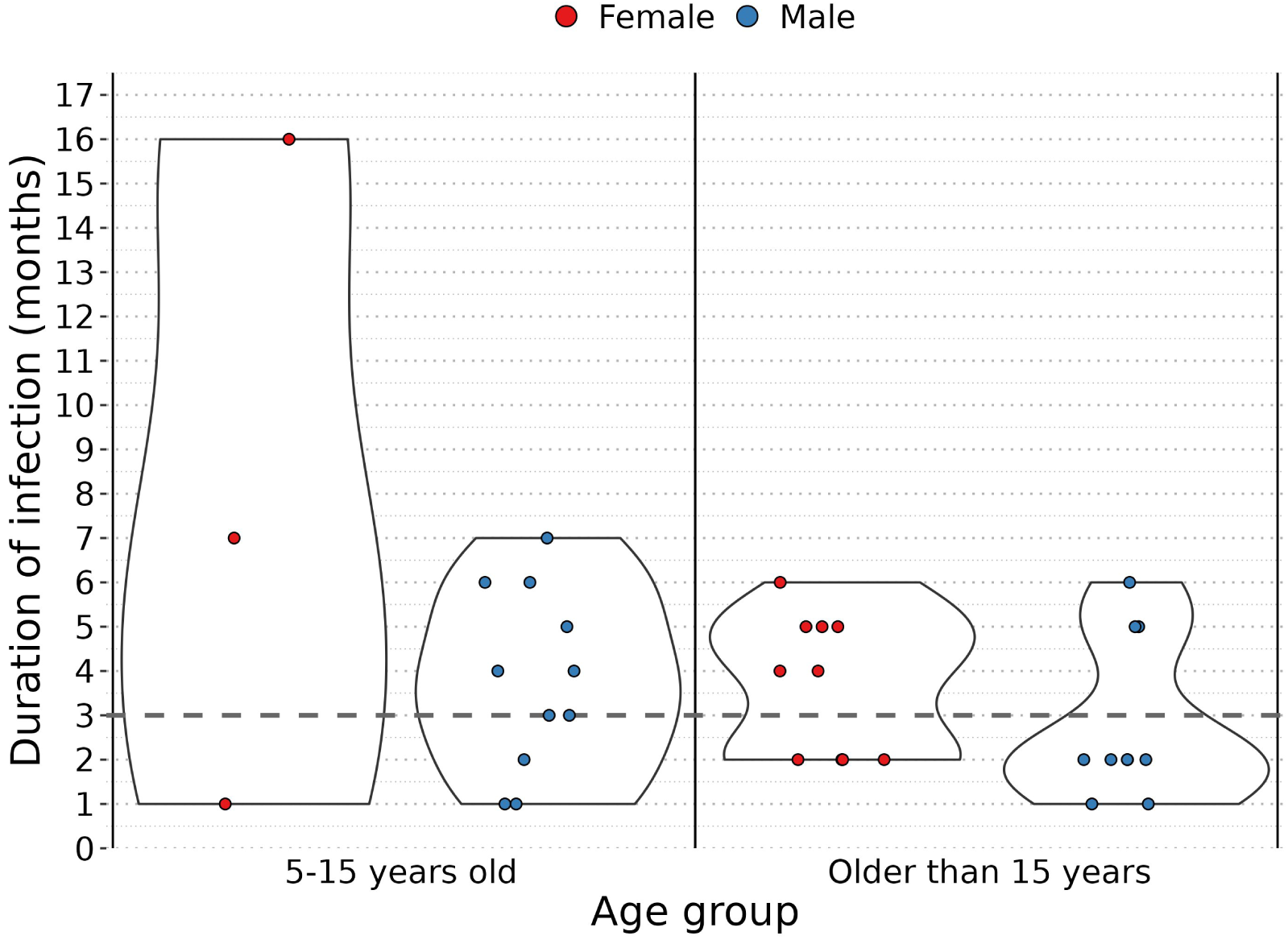
Durations of infection by the same parasite strain, grouped by host age and coloured by gender. The number of short (less than 3 months) and long infections (more than 3 months) were compared between genders (female versus male) and age groups (‘5-15 years old’ versus ‘older than 15 years’). No significant difference of the duration of infection was found between genders, which had very similar percentages of short infections with 38 % and 48 % of infections for female and male respectively (χ^2^ = 0.30, p-value > 0.5). Although long infections were more common in the ‘5-15 years old’ age group (71 % of all infections) compared to the ‘older than 15 years age’ group (45 % of all infections), this was not statistically significant (χ^2^ = 2.34, p-value < 0.13), potentially due to relatively small sample size.

## References

1. World malaria report 2023. 13 Mar 2024 [cited 13 Mar 2024]. Available: https://www.who.int/teams/global-malaria-programme/reports/world-malaria-report-2023

2. Ceesay SJ, Casals-Pascual C, Erskine J, Anya SE, Duah NO, Fulford AJ, et al. Changes in malaria indices between 1999 and 2007 in The Gambia: a retrospective analysis. The Lancet. 2008;372: 1545–1554. doi:10.1016/S0140-6736(08)61654-2

3. Mwesigwa J, Okebe J, Affara M, Di Tanna GL, Nwakanma D, Janha O, et al. On-going malaria transmission in The Gambia despite high coverage of control interventions: a nationwide cross-sectional survey. Malar J. 2015;14: 314. doi:10.1186/s12936-015-0829-6

4. Mwesigwa J, Achan J, Tanna GLD, Affara M, Jawara M, Worwui A, et al. Residual malaria transmission dynamics varies across The Gambia despite high coverage of control interventions. PLOS ONE. 2017;12: e0187059. doi:10.1371/journal.pone.0187059

5. Sonko ST, Jaiteh M, Jafali J, Jarju LB, D’Alessandro U, Camara A, et al. Does socio-economic status explain the differentials in malaria parasite prevalence? Evidence from The Gambia. Malar J. 2014;13: 449. doi:10.1186/1475-2875-13-449

6. Ahmad A, Mohammed NI, Joof F, Affara M, Jawara M, Abubakar I, et al. Asymptomatic Plasmodium falciparum carriage and clinical disease: a 5-year community-based longitudinal study in The Gambia. Malar J. 2023;22: 82. doi:10.1186/s12936-023-04519-0

7. Fola AA, Moser KA, Aydemir O, Hennelly C, Kobayashi T, Shields T, et al. Temporal and spatial analysis of Plasmodium falciparum genomics reveals patterns of parasite connectivity in a low-transmission district in Southern Province, Zambia. Malar J. 2023;22: 208. doi:10.1186/s12936-023-04637-9

8. Gwarinda HB, Tessema SK, Raman J, Greenhouse B, Birkholtz L-M. Parasite genetic diversity reflects continued residual malaria transmission in Vhembe District, a hotspot in the Limpopo Province of South Africa. Malar J. 2021;20: 96. doi:10.1186/s12936-021-03635-z

9. Sy M, Deme AB, Warren JL, Early A, Schaffner S, Daniels RF, et al. Plasmodium falciparum genomic surveillance reveals spatial and temporal trends, association of genetic and physical distance, and household clustering. Sci Rep. 2022;12: 938. doi:10.1038/s41598-021-04572-2

10. Daniels RF, Schaffner SF, Wenger EA, Proctor JL, Chang H-H, Wong W, et al. Modeling malaria genomics reveals transmission decline and rebound in Senegal. Proc Natl Acad Sci. 2015;112: 7067–7072. doi:10.1073/pnas.1505691112

11. Collins KA, Ceesay S, Drammeh S, Jaiteh FK, Guery MA, Lanke K, et al. A Cohort Study on the Duration of Plasmodium falciparum Infections During the Dry Season in The Gambia. J Infect Dis. 2022;226: 128–137. doi:10.1093/infdis/jiac116

12. Andolina C, Rek JC, Briggs J, Okoth J, Musiime A, Ramjith J, et al. Sources of persistent malaria transmission in a setting with effective malaria control in eastern Uganda: a longitudinal, observational cohort study. Lancet Infect Dis. 2021;21: 1568–1578. doi:10.1016/S1473-3099(21)00072-4

13. Barry A, Bradley J, Stone W, Guelbeogo MW, Lanke K, Ouedraogo A, et al. Higher gametocyte production and mosquito infectivity in chronic compared to incident Plasmodium falciparum infections. Nat Commun. 2021;12: 2443. doi:10.1038/s41467-021-22573-7

14. Nkhoma SC, Nair S, Cheeseman IH, Rohr-Allegrini C, Singlam S, Nosten F, et al. Close kinship within multiple-genotype malaria parasite infections. Proc R Soc B Biol Sci. 2012;279: 2589–2598. doi:10.1098/rspb.2012.0113

15. Daniels R, Chang H-H, Séne PD, Park DC, Neafsey DE, Schaffner SF, et al. Genetic Surveillance Detects Both Clonal and Epidemic Transmission of Malaria following Enhanced Intervention in Senegal. Mayor A, editor. PLoS ONE. 2013;8: e60780. doi:10.1371/journal.pone.0060780

16. Pacheco MA, Forero-Peña DA, Schneider KA, Chavero M, Gamardo A, Figuera L, et al. Malaria in Venezuela: changes in the complexity of infection reflects the increment in transmission intensity. Malar J. 2020;19: 176. doi:10.1186/s12936-020-03247-z

17. Hendry JA, Kwiatkowski D, McVean G. Elucidating relationships between P.falciparum prevalence and measures of genetic diversity with a combined genetic-epidemiological model of malaria. PLoS Comput Biol. 2021;17: e1009287. doi:10.1371/journal.pcbi.1009287

18. Lee A, Ndiaye YD, Badiane A, Deme A, Daniels RF, Schaffner SF, et al. Modeling the levels, trends, and connectivity of malaria transmission using genomic data from a health facility in Thiès, Senegal. medRxiv; 2021 Oct p. 2021.09.17.21263639. doi:10.1101/2021.09.17.21263639

19. Daniels R, Volkman SK, Milner DA, Mahesh N, Neafsey DE, Park DJ, et al. A general SNP-based molecular barcode for Plasmodium falciparum identification and tracking. Malar J. 2008;7: 223. doi:10.1186/1475-2875-7-223

20. Amambua-Ngwa A, Jeffries D, Mwesigwa J, Seedy-Jawara A, Okebe J, Achan J, et al. Long-distance transmission patterns modelled from SNP barcodes of Plasmodium falciparum infections in The Gambia. Sci Rep. 2019;9: 13515. doi:10.1038/s41598-019-49991-4

21. Mobegi VA, Loua KM, Ahouidi AD, Satoguina J, Nwakanma DC, Amambua-Ngwa A, et al. Population genetic structure of Plasmodium falciparum across a region of diverse endemicity in West Africa. Malar J. 2012;11: 223. doi:10.1186/1475-2875-11-223

22. Taylor AR, Schaffner SF, Cerqueira GC, Nkhoma SC, Anderson TJC, Sriprawat K, et al. Quantifying connectivity between local Plasmodium falciparum malaria parasite populations using identity by descent. Didelot X, editor. PLOS Genet. 2017;13: e1007065. doi:10.1371/journal.pgen.1007065

23. Noviyanti R, Miotto O, Barry A, Marfurt J, Siegel S, Thuy-Nhien N, et al. Implementing parasite genotyping into national surveillance frameworks: feedback from control programmes and researchers in the Asia–Pacific region. Malar J. 2020;19: 271. doi:10.1186/s12936-020-03330-5

24. Lindblade KA, Steinhardt L, Samuels A, Kachur SP, Slutsker L. The silent threat: asymptomatic parasitemia and malaria transmission. Expert Rev Anti Infect Ther. 2013;11: 623–639. doi:10.1586/eri.13.45

25. Stone W, Gonçalves BP, Bousema T, Drakeley C. Assessing the infectious reservoir of falciparum malaria: past and future. Trends Parasitol. 2015;31: 287–296. doi:10.1016/j.pt.2015.04.004

26. Fogang B, Lellouche L, Ceesay S, Drammeh S, Jaiteh FK, Guery M-A, et al. Asymptomatic Plasmodium falciparum carriage at the end of the dry season is associated with subsequent infection and clinical malaria in Eastern Gambia. Malar J. 2024;23: 22. doi:10.1186/s12936-024-04836-y

27. Jacob CG, Thuy-Nhien N, Mayxay M, Maude RJ, Quang HH, Hongvanthong B, et al. Genetic surveillance in the Greater Mekong subregion and South Asia to support malaria control and elimination. Neafsey DE, Soldati-Favre D, Neafsey DE, Ménard D, editors. eLife. 2021;10: e62997. doi:10.7554/eLife.62997

28. Oyola SO, Ariani CV, Hamilton WL, Kekre M, Amenga-Etego LN, Ghansah A, et al. Whole genome sequencing of Plasmodium falciparum from dried blood spots using selective whole genome amplification. Malar J. 2016;15: 597. doi:10.1186/s12936-016-1641-7

29. McKenna A, Hanna M, Banks E, Sivachenko A, Cibulskis K, Kernytsky A, et al. The Genome Analysis Toolkit: A MapReduce framework for analyzing next-generation DNA sequencing data. Genome Res. 2010;20: 1297–1303. doi:10.1101/gr.107524.110

30. Malaria GEN, Ahouidi A, Ali M, Almagro-Garcia J, Amambua-Ngwa A, Amaratunga C, et al. An open dataset of Plasmodium falciparum genome variation in 7,000 worldwide samples. Wellcome Open Res. 2021;6: 42. doi:10.12688/wellcomeopenres.16168.2

31. Schaffner SF, Taylor AR, Wong W, Wirth DF, Neafsey DE. hmmIBD: software to infer pairwise identity by descent between haploid genotypes. Malar J. 2018;17: 196. doi:10.1186/s12936-018-2349-7

32. Miles A, Iqbal Z, Vauterin P, Pearson R, Campino S, Theron M, et al. Indels, structural variation, and recombination drive genomic diversity in *Plasmodium* falciparum. Genome Res. 2016;26: 1288–1299. doi:10.1101/gr.203711.115

33. Manske M, Miotto O, Campino S, Auburn S, Almagro-Garcia J, Maslen G, et al. Analysis of Plasmodium falciparum diversity in natural infections by deep sequencing. Nature. 2012;487: 375–379. doi:10.1038/nature11174

34. McHugh ML. Interrater reliability: the kappa statistic. Biochem Medica. 2012;22: 276.

35. Nwakanma DC, Duffy CW, Amambua-Ngwa A, Oriero EC, Bojang KA, Pinder M, et al. Changes in Malaria Parasite Drug Resistance in an Endemic Population Over a 25-Year Period With Resulting Genomic Evidence of Selection. J Infect Dis. 2014;209: 1126–1135. doi:10.1093/infdis/jit618

36. World Health Organization. WHO policy recommendation: seasonal malaria chemoprevention (SMC) for plasmodium falciparum malaria control in highly seasonal transmission areas of the Sahel sub-region in Africa. 2012 [cited 15 Jan 2025]. Available: https://iris.who.int/handle/10665/337978

37. Echeverry DF, Nair S, Osorio L, Menon S, Murillo C, Anderson TJ. Long term persistence of clonal malaria parasite Plasmodium falciparum lineages in the Colombian Pacific region. BMC Genet. 2013;14: 2. doi:10.1186/1471-2156-14-2

38. Taylor AR, Jacob PE, Neafsey DE, Buckee CO. Estimating Relatedness Between Malaria Parasites. Genetics. 2019;212: 1337–1351. doi:10.1534/genetics.119.302120

39. Redmond SN, MacInnis BM, Bopp S, Bei AK, Ndiaye D, Hartl DL, et al. De Novo Mutations Resolve Disease Transmission Pathways in Clonal Malaria. Mol Biol Evol. 2018;35: 1678–1689. doi:10.1093/molbev/msy059

40. Camponovo F, Buckee CO, Taylor AR. Measurably recombining malaria parasites. Trends Parasitol. 2023;39: 17–25. doi:10.1016/j.pt.2022.11.002

41. Nkhoma SC, Trevino SG, Gorena KM, Nair S, Khoswe S, Jett C, et al. Co-transmission of Related Malaria Parasite Lineages Shapes Within-Host Parasite Diversity. Cell Host Microbe. 2020;27: 93–103.e4. doi:10.1016/j.chom.2019.12.001

42. Zhu SJ, Almagro-Garcia J, McVean G. Deconvolution of multiple infections in Plasmodium falciparum from high throughput sequencing data. Birol I, editor. Bioinformatics. 2018;34: 9–15. doi:10.1093/bioinformatics/btx530

43. Taylor AR, Echeverry DF, Anderson TJC, Neafsey DE, Buckee CO. Identity-by-descent with uncertainty characterises connectivity of Plasmodium falciparum populations on the Colombian-Pacific coast. PLOS Genet. 2020;16: e1009101. doi:10.1371/journal.pgen.1009101

44. Okell LC, Bousema T, Griffin JT, Ouédraogo AL, Ghani AC, Drakeley CJ. Factors determining the occurrence of submicroscopic malaria infections and their relevance for control. Nat Commun. 2012;3: 1237. doi:10.1038/ncomms2241

45. Slater HC, Ross A, Felger I, Hofmann NE, Robinson L, Cook J, et al. The temporal dynamics and infectiousness of subpatent Plasmodium falciparum infections in relation to parasite density. Nat Commun. 2019;10: 1433. doi:10.1038/s41467-019-09441-1

46. Ashley EA, White NJ. The duration of Plasmodium falciparum infections. Malar J. 2014;13: 500. doi:10.1186/1475-2875-13-500

47. Bylicka-Szczepanowska E, Korzeniewski K. Asymptomatic Malaria Infections in the Time of COVID-19 Pandemic: Experience from the Central African Republic. Int J Environ Res Public Health. 2022;19: 3544. doi:10.3390/ijerph19063544

48. Mueller I, Schoepflin S, Smith TA, Benton KL, Bretscher MT, Lin E, et al. Force of infection is key to understanding the epidemiology of Plasmodium falciparum malaria in Papua New Guinean children. Proc Natl Acad Sci U S A. 2012;109: 10030–10035. doi:10.1073/pnas.1200841109

49. Conway DJ, McBride JS. Genetic evidence for the importance of interrupted feeding by mosquitoes in the transmission of malaria. Trans R Soc Trop Med Hyg. 1991;85: 454–456. doi:10.1016/0035-9203(91)90217-M

50. Steinhardt LC, Kc A, Tiffany A, Quincer EM, Loerinc L, Laramee N, et al. Reactive Case Detection and Treatment and Reactive Drug Administration for Reducing Malaria Transmission: A Systematic Review and Meta-Analysis. Am J Trop Med Hyg. 2024;110: 82–93. doi:10.4269/ajtmh.22-0720

51. Newby G, Cotter C, Roh ME, Harvard K, Bennett A, Hwang J, et al. Testing and treatment for malaria elimination: a systematic review. Malar J. 2023;22: 254. doi:10.1186/s12936-023-04670-8

52. Stresman G, Whittaker C, Slater HC, Bousema T, Cook J. Quantifying Plasmodium falciparum infections clustering within households to inform household-based intervention strategies for malaria control programs: An observational study and meta-analysis from 41 malaria-endemic countries. PLoS Med. 2020;17: e1003370. doi:10.1371/journal.pmed.1003370

53. Okebe J, Dabira E, Jaiteh F, Mohammed N, Bradley J, Drammeh N-F, et al. Reactive, self-administered malaria treatment against asymptomatic malaria infection: results of a cluster randomized controlled trial in The Gambia. Malar J. 2021;20: 253. doi:10.1186/s12936-021-03761-8

54. Soremekun S, Conteh B, Nyassi A, Soumare HM, Etoketim B, Ndiath MO, et al. Household-level effects of seasonal malaria chemoprevention in the Gambia. Commun Med. 2024;4: 1–11. doi:10.1038/s43856-024-00503-0

